# Modeling of aerosol transmission of airborne pathogens in ICU rooms of COVID-19 patients with acute respiratory failure

**DOI:** 10.1101/2020.12.11.20247551

**Authors:** Cyril Crawford, Emmanuel Vanoli, Baptiste Decorde, Maxime Lancelot, Camille Duprat, Christophe Josserand, Jonathan Jilesen, Lila Bouadma, Jean-François Timsit

## Abstract

The COVID-19 pandemic has generated many concerns about cross-contamination risks, particularly in hospital settings and Intensive Care Units (ICU). Virus-laden aerosols produced by infected patients can propagate throughout ventilated rooms and put medical personnel entering them at risk. Experimental results found with a schlieren optical method have shown that the air flows generated by a cough and normal breathing were modified by the oxygenation technique used, especially when using High Flow Nasal Canulae, increasing the shedding of potentially infectious airborne particles. This study also uses a 3D Computational Fluid Dynamics model based on a Lattice Boltzmann Method to simulate the air flows as well as the movement of numerous airborne particles produced by a patient’s cough within an ICU room under negative pressure. The effects of different mitigation scenarii on the amount of aerosols potentially containing SARS-CoV-2 that are extracted through the ventilation system are investigated. Numerical results indicate that adequate bed orientation and additional air treatment unit positioning can increase by 40% the number of particles extracted and decrease by 25% the amount of particles deposited on surfaces 45s after shedding. This approach could help lay the grounds for a more comprehensive way to tackle contamination risks in hospitals, as the model can be seen as a proof of concept and be adapted to any room configuration.

## Introduction

The World Health Organisation (WHO) has reported that the recent pandemic of SARS-CoV-2 (COVID-19), which has led to over 135 million persons being contaminated worldwide^1^, can be propagated through various means. One of the main transmission vectors is assumed to be ‘respiratory droplets’ (diameter over 5 *µ*m) and fomites. However, aerosol transmission via ‘droplet nuclei’ (diameter under 5 *µ*m) is also being considered in order to explain high infection rates, although its relative importance remains controversial: this type of particle does not sediment quickly and follows airflows inertially^2–4^. Aerosol transmission risk has been studied for decades, with influenza for example^5^. When breathing, sneezing or coughing, many particles with a diameter comprised between 100 nm and 1 mm are excreted at speeds ranging from 1 to 10 m/s, with some being small enough to be inhaled deeply in the human respiratory tract^6^. They can travel up to 10 meters in indoor spaces and are of particular concern^7,8^. Additionally, viral particles can remain airborne and infectious for hours in an artificially-created aerosol^9^, emphasizing the importance of the risk of infection in indoor environments. While airborne transmission has been suggested in a Wuhan restaurant with poor ventilation^10^, the risk is even more acute for healthcare professionals who work in a hospital environment where COVID-19 patients are being treated^11^. The virus has infected more people who work in hospital settings on average than in the general population, as seroprevalence studies and meta-analyses have shown^12–15^. However, this claim has been disputed, since medical personnel wear Personal Protective Equipment that shields them more efficiently from the virus than the general population^16,17^. Nevertheless, it is essential to find ways to mitigate risks for these frontline workers. Furthermore, the number of hospitalized COVID-19 patients that are sent to Intensive Care Units (ICU) for treatment ranges between 17% and 35%^18^. Many medical procedures in ICUs have a high potential for aerosol generation, putting medical staff at even higher risk of exposure to the virus^19^. These methods include endotracheal intubation, an intrusive procedure which precedes mechanical ventilation. It has been used extensively to treat COVID-19 patients in critical state and is associated with high mortality^20^. Controversy exists regarding the threshold for intubation because many patients with severe hypoxemia display a normal work of breathing, without any distress: early intubation lowers the risk of contamination of the medical personnel and is logistically simpler, but might be useless and only add complications for the patient^18^. Clinicians must be careful when assessing a patient’s need for intubation because of the short supply of ventilators and the underlying risk of complications linked to sedation and muscle paralysis^21,22^. Lessons from the MERS-CoV epidemic have indicated that less intrusive oxygenation techniques were to be avoided due to increased risk of contamination through droplets^23^. However, these less harmful methods are increasingly being used to treat COVID-19 patients in ICUs and can be divided among the following three categories: conventional Noninvasive Ventilation (NIV), High Flow Nasal Canulae (HFNC), and Continuous Positive Airway Pressure (CPAP)^24^. The problem associated with these types of mechanically assisted ventilation is that oxygen is supplied at rates that can be as high as 60L/min for HFNC, thus increasing the volume and reach of exhaled air^25^. Despite higher airborne contamination risks for healthcare workers working in the rooms of patients under Noninvasive Ventilation^26^, they have been associated with improved patient outcomes^27^, although this claim is subject to debate^22,28^. Moreover, the necessary presence of Heating Ventilation and Air-Conditioning (HVAC) systems providing fresh air in hospital rooms may also play a key role in the risk of airborne infection: this is emphasized in the Wells-Riley equation which links the probability of indoors airborne transmission of pathogens not only to the volume flux of air exhaled by an infected patient but also the volume flux of clean air injected in the room by the HVAC^29^. Displacement ventilation can reduce transmission risk as opposed to mixing ventilation, however this knowledge is not useful for rooms where HVAC is already installed^29^. It is therefore necessary to assess the risk for medical workers and to explore simple mitigating scenarii.

The purpose of our work is to model precisely the movement of aerosols produced by a patient with HFNC in an ICU room. Once the patterns of extraction and deposition of these potentially virus-laden particles have been successfully compared under different room configurations, measures can be proposed to mitigate the risk of transmission by maximizing the number of extracted particles through ventilation and optimizing the cleaning of relevant surfaces. Although each room has its own specificities, our numerical modelling approach can be adapted on a case by case basis to any hospital room and can thus be seen as a proof of concept. Several numerical modeling attempts of aerosol propagation within a ventilated hospital room have been made over the past few years. Most are too time-consuming and not precise enough to be able to yield useful and generalizable advice for hospital staff. Nevertheless, Computational Fluid Dynamics (CFD) simulations have already been used previously to assess the propagation dynamics of infectious aerosols in an indoor ventilated environment. A first CFD study concluded that the ventilation type and the position of the inlet and outlet were crucial elements to be considered^30^. However these results are only useful when designing a new room and do not provide a way to optimize the configuration of a room that is already built. A second CFD model using a Langrangian Stochastic flight model was able to calculate the propagation of aerosols over time and showed the influence of the source’s height^31^. Unfortunately, this model uses probability distribution and the analysis are unidimensional, making it impractical. The dimensionality issue was partly solved in a subsequent study, which proposed a 3D CFD model^32^. Finally, CFD models have been used more recently to describe the dynamics of aerosols within a car^33^ as well as those being ejected during a violent expiratory event, showing that wind could help propagate them over distances that can reach 6m^34^. These models was not used to optimize hospital room configuration. Some models have also been used to predict the inactivation of airborne pathogens by Ultraviolet C lighting^35^. Additionally, other CFD models were used to predict zones of high exposure to aerosols and link them to areas where infection patterns had been reported in the real-life setting of a Wuhan restaurant^36^. Lagrangian statistics have been used to demonstrate that small droplets’ lifetime is longer than previously expected due to the turbulent humid air expelled with them when breathing or coughing, resulting in increased potential propagation distances^37^. Overall, these models emphasize the impact of room conditions, source height as well as position and ventilation type on the propagation of potentially infected aerosols.

The first set of factors to consider when looking at the production of aerosols by infected patients is the size distribution and number of particles emitted during a respiratory event. They can be measured through spectrometry: Laser Particle Spectrometry^38^, Aerodynamic Particle Sizer^39^ and Scanning Mobility Particle Sizer^40^. The size distribution commonly considered for normally paced breathing is log-normal, with a peak centered around 1 *µ*m or lower^39,41,42^. This peak is located around the same value for more violent events (talking, singing, coughing)^39^. Other sources present higher values^34,43,44^ but evaporation and hygroscopy have to be taken into account^45,46^. The associated reduction in size of the droplets occurs very fast: less than a second for droplets under 10 *µ*m^46,47^. The evaporation time depends on the temperature and relative humidity of the room^43^: the aerosol particles followed throughout the room during a time interval spanning several minutes are thus fully reduced to their droplet nuclei. Furthermore, the average number of particles that are exhaled by a patient breathing normally is around 1000 per second and can go up to 10000 per second when coughing^48^. However, more advanced measurement techniques have yielded more accurate results: 4 × 10^5^ un-evaporated droplets are emitted per second in the 0.6-40 *µ*m size range for coughing^41^. The size of particles containing virions also has an impact on how deep these can penetrate the respiratory tract and find mucous membrane that are more easily penetrable^49^. The saliva composition can also play a role in the model, as well as the probability of presence of virus in a droplet. It is indeed important to note that not all particles exhaled by an infected person contain pathogens. Furthermore it is still unclear how the shedding of aerosolized pathogens result in infection, as the minimal inhaled viral load required to infect someone is unknown. The flow rate of the patient’s exhalation is another essential parameter to take into consideration and needs to be measured for different breathing apparatus. A schlieren optical technique can be adequately used to study the dynamics of fluids and gases and can thus be applied to characterize exhaled air. The second set of parameters to take into account relates to the morphology of the room, which is under negative pressure: dimensions, patient position, intensity of room depression, ventilation parameters, additional air treatment unit position. These parameters are described in the Methods section. The room we chose to represent is a typical single occupancy one and is representative of those found in hospitals and in ICUs in particular. Its HVAC system has an outlet in the corner left on the left side of the patient and an inlet above him, in the center of the room (Supplementary Fig. S6). The goal is to test different room configurations (bed placement, ventilation mode, position of an additional air treatment unit) and to determine the optimal one in terms of reduced risk of infection. Four configurations were studied and are presented in Fig. 1. They correspond to different use cases: the last two configurations present a mobile air treatment unit positioned on the axis that links the HVAC’s inlet vent to its outlet vent, on either side of the patient’s bed. The first two configurations do not have this additional unit: only the bed orientation varies.

**Figure 1.**
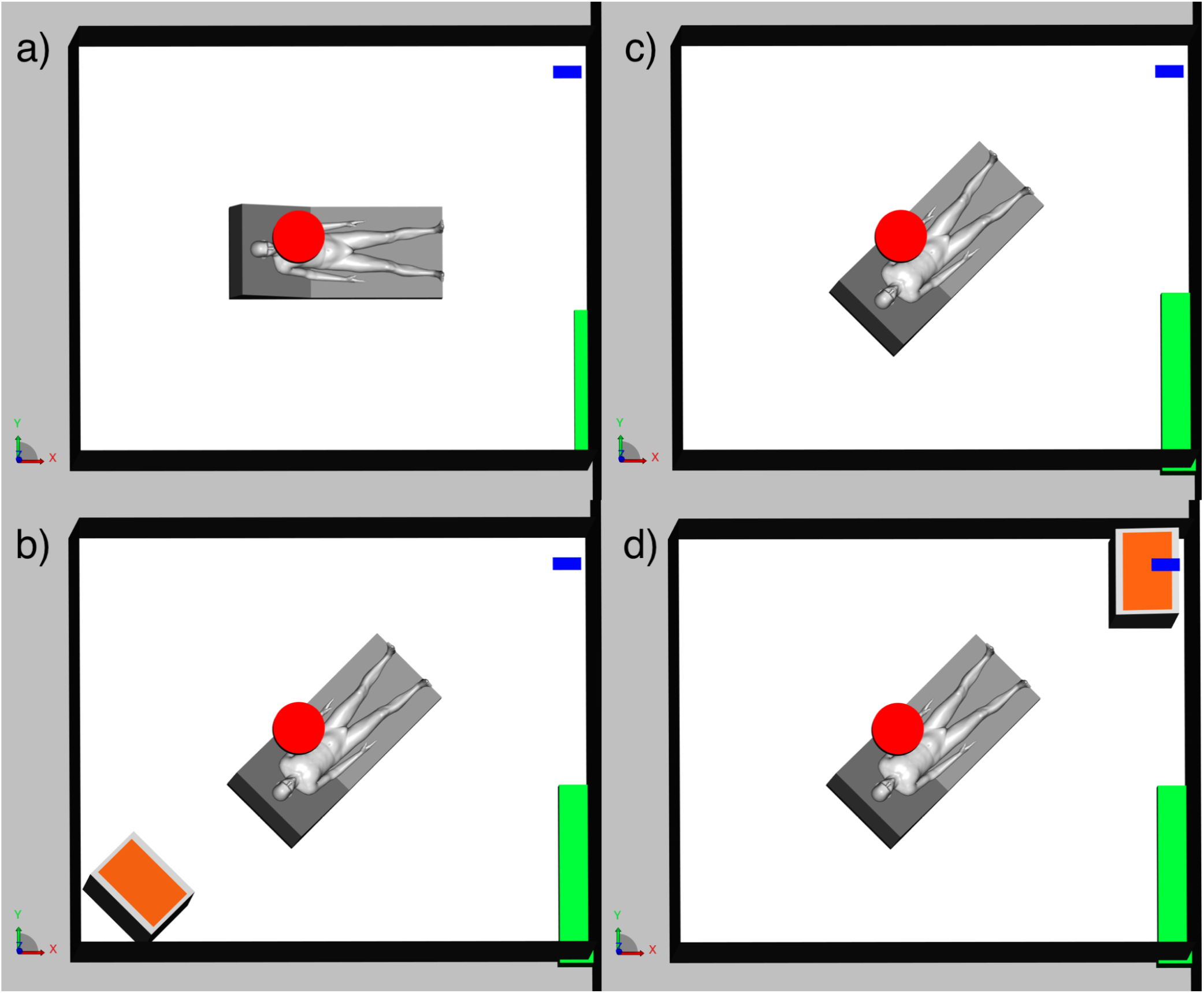
Room configuration for the 4 scenarios that were modelled: (a) Baseline configuration; (c) 45° bed orientation configuration; (b) Plasmair + 45° bed orientation in configuration 1; (d) Plasmair + 45° bed orientation in configuration 2. The ventilation’s inlet vent is represented in red, and it’s outlet vent is in blue. The gap under the door is in green and the Plasmair is in orange.

## Results

The purpose of the optical experiments is to calibrate the numerical model and to ensure that it is as precise and realistic as possible. We use an ombroscopy technique to obtain high speed visualizations and flow field measurements of the exhaled air (breathing or coughing) of healthy volunteers with different breathing apparatus. The results of the high-resolution video of a cough made by a volunteer using HFNC at 60L/min are shown in Fig. 2. The characteristic propagation cone is visible in the view from above (Fig. 2a) and its angle can reach 70°. The turbulent eddies of the jet produced by this respiratory event as well as the jet front are clearly visible in the side view (Fig. 2b). The Particle Image Velocimetry (PIV)-analyzed vector map in Fig. 2c indicates that the maximum average air speed is around 5 m/s and is located at the turbulent front. Flow rates can reach up to 4 L/s. The higher the oxygen input flow rate, the higher the exhaled flow rate is when breathing (Supplementary Fig. S4 and S5). Other schlieren experiments were done on volunteers using the CPAP oxygenation apparatus. Breathing regimes were analyzed, with oxygen input rates of 15L/min and 30L/min. Results are also presented in Fig. 2. Leaks occur at the interface between the mask and the volunteer’s face and are located around the top of the nose, oriented upwards (Fig. 2d) and on the cheeks, oriented sideways (Fig. 2e). A qualitative analysis indicates that the higher the oxygen input flow rate, the more substantial the leaks are. The flow rates of these leaks and local air velocities are very small compared to those produced by HFNC, as shown on the vector map of Fig. 2f, where the maximum average air speed is around 0.4 m/s. Potential aerosol propagation within the room is thus hindered when using CPAP over HFNC and when lowering the flow of input oxygen. CPAP results were not used for the numerical modeling because the leaks were found to be negligible compared to air flows induced by HFNC. Complete films of the experiments are provided in the Electronic Supplement Materials, as well as flow rate analysis (Supplementary Fig. S4 and S5).

**Figure 2.**
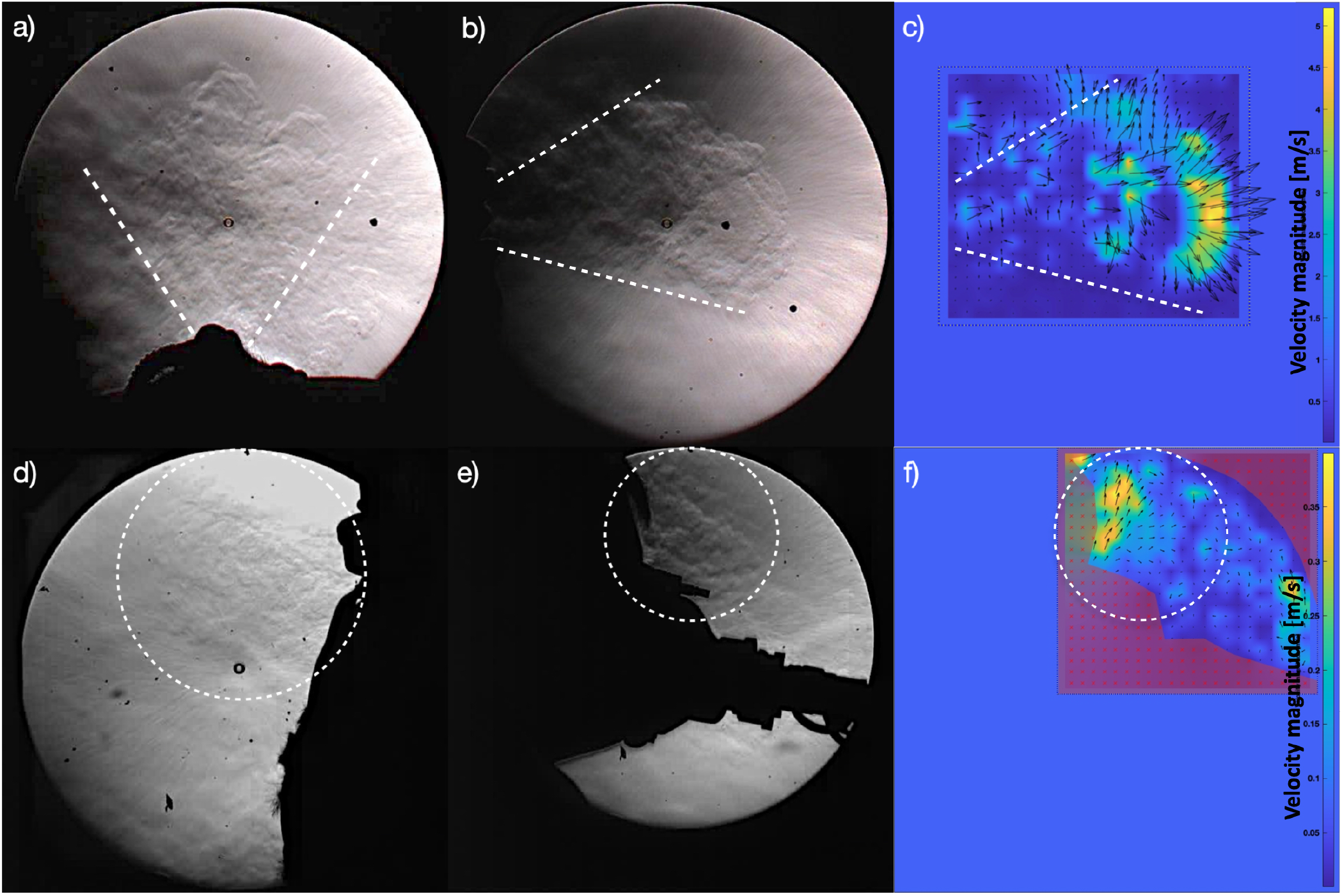
Qualitative and quantitative results from schlieren images of a volunteer’s cough with High Flow Nasal Canulae (HFNC) at 60L/min (top) and with Continuous Positive Airway Pressure (CPAP) at 30L/min (bottom): (a) & (d) View from above with the mouth on the bottom side; (b) & (e) Side view with the mouth on the left side; (c) & (f) Instantaneous velocity magnitude vectors collected through Particle Image Velocimetry (PIV) processing of the ‘side view’ video frames. For HFNC, there are exhaled air speed maximas of 5 m/s at the wave front. For CPAP, leaked air speed maximas reach 0.4 m/s. Air flows, and thus aerosol shedding, depend on the oxygenation technique: they are much more extensive with HFNC than with CPAP. Video sequences are provided in the supplementary material.

The simulation model was run with a patient equipped with HFNC coughing in an ICU room under negative pressure. To validate it and make sure that the complexity of such a flow is coherently represented, the images obtained from the simulation are compared to the schlieren images of the physical tests at different key phases of the cough. Shortly after the start of the cough, (65 ms), a jet and a “mushroom” start to develop similarly downstream of the mouth for both the test and the simulation (Fig. 3a). Later on, after 150 ms, while the jet is still quite straight from the upper side of the mouth, it slopes downward from the lower lip. In both cases, a “bump” in the density gradient can be seen, (Fig. 3b). Finally, after 190 ms, the position of the jet fluctuations in the test and in the simulation are still similar, as well as the spatial expansion of the jet, despite the HFNC system providing additional fluctuations at the top of the jet (Fig. 3c). The modeled mouth geometry is not a scan of the test person, hence there seems to be limited dependency between the pattern of the jet and the shape of the mouth internals.

**Figure 3.**
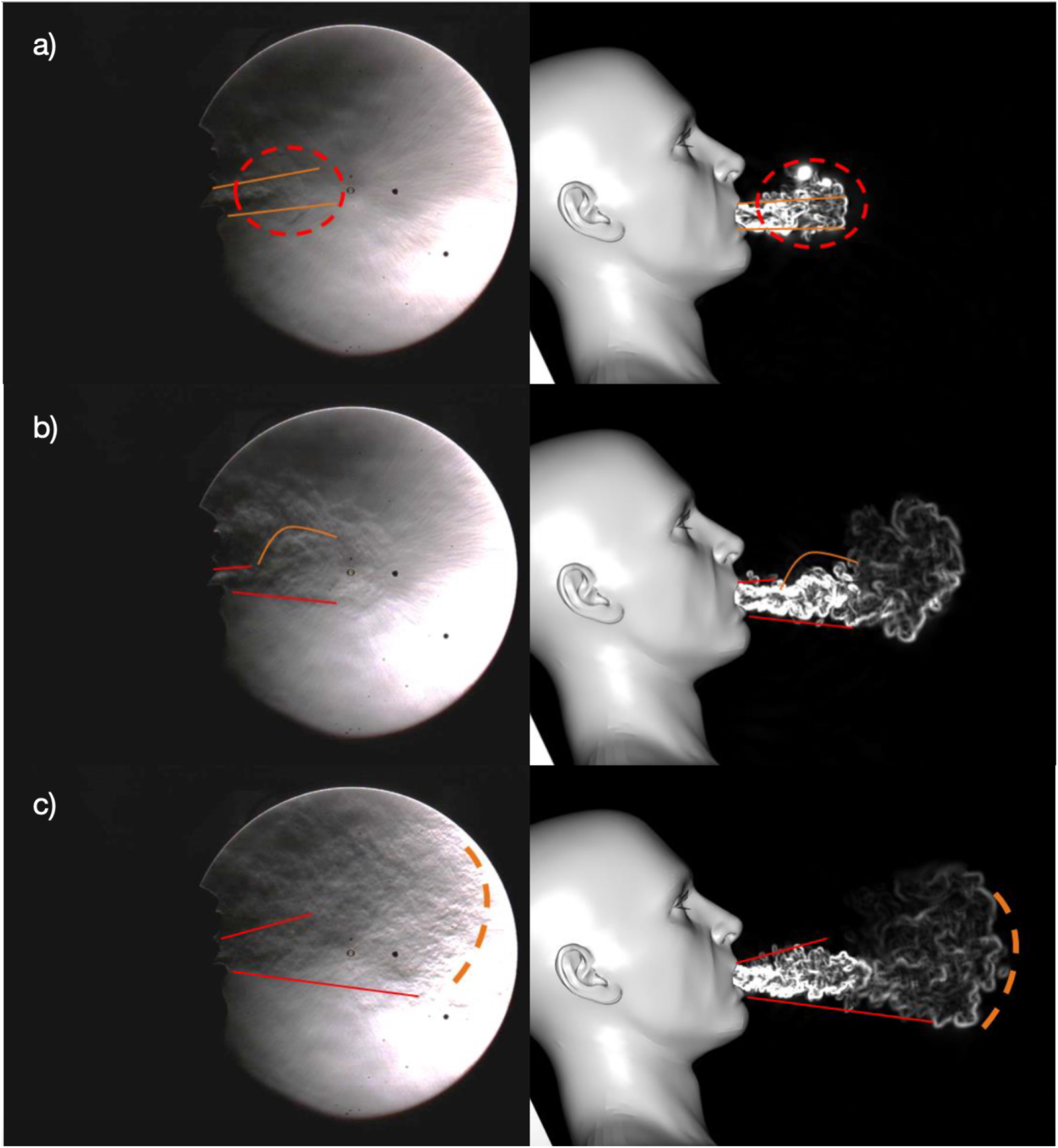
Air density gradient visualization of a volunteer’s cough with HFNC at 60L/min taken from the schlieren images (left) and the associated numerical simulations (right): (a) at t = 65ms; (b) at t = 150ms; (c) at t = 190ms. A good correlation is observed between the experiment and the simulation for the front and the bottom part of the jet. Leaks coming from the nose can explain the differences observed for the upper part of the jet.

With the first phase validated, airflows and particle dispersion were then simulated within the room. Two scenarios were investigated first: a baseline configuration where the room is in a normal state (Fig. 1a); and a 45° configuration in which the bed is oriented towards the ventilation outlet (Fig. 1c). Two additionnal scenarios were tested afterwards, with an additional mobile air treatment unit (Plasmair) placed on the axis between the ventilation outlet vent and the patient’s bed turned at 45°: a first configuration where the Plasmair is behind the bed (Fig. 1b); and a second where it is located in front of the bed and under the outlet (Fig. 1d).

Due to the morphology of the room and ventilation system, the air flow around the patient is oriented towards the ceiling. The impact of the negative pressure is highlighted by a high velocity air jet coming inside the room from under the door (see Supplementary Fig. S7 online). It leads to a turbulent flow around the patient which is oriented towards the ceiling due to the ventilation system (Supplementary Fig. S8). Furthermore, body temperature is higher than room temperature and hot air tends to rise upwards. The exhaled particles will follow these two trends, as seen in Fig. 4a, and quickly reach the ceiling area where the ventilation system’s inlet is located. This type of ventilation, common in hospital settings, dispatches the air at 360° within a plane parallel to the ceiling. As soon as the particles reach the surroundings of the inlet, they follow the air flows and are dispatched in all directions, although most are directed towards the outlet as shown in Fig. 4b. This strong dispersion increases the risk of shedding potentially infected airborne particles all over the room. It also increases their deposit on surfaces (walls, tables, ceiling, medical equipment…). The blue points of impact of particles on the surfaces after 45s of physical simulated time presented in Fig. 4c highlight a high deposit concentration on the side where the air extraction is positioned. Impact points are present on a large portion of the floor, whereas the bed is much less impacted by these small particles. This is due to the air moving up quickly to the ceiling: particles do not remain airborne around the patient.

**Figure 4.**
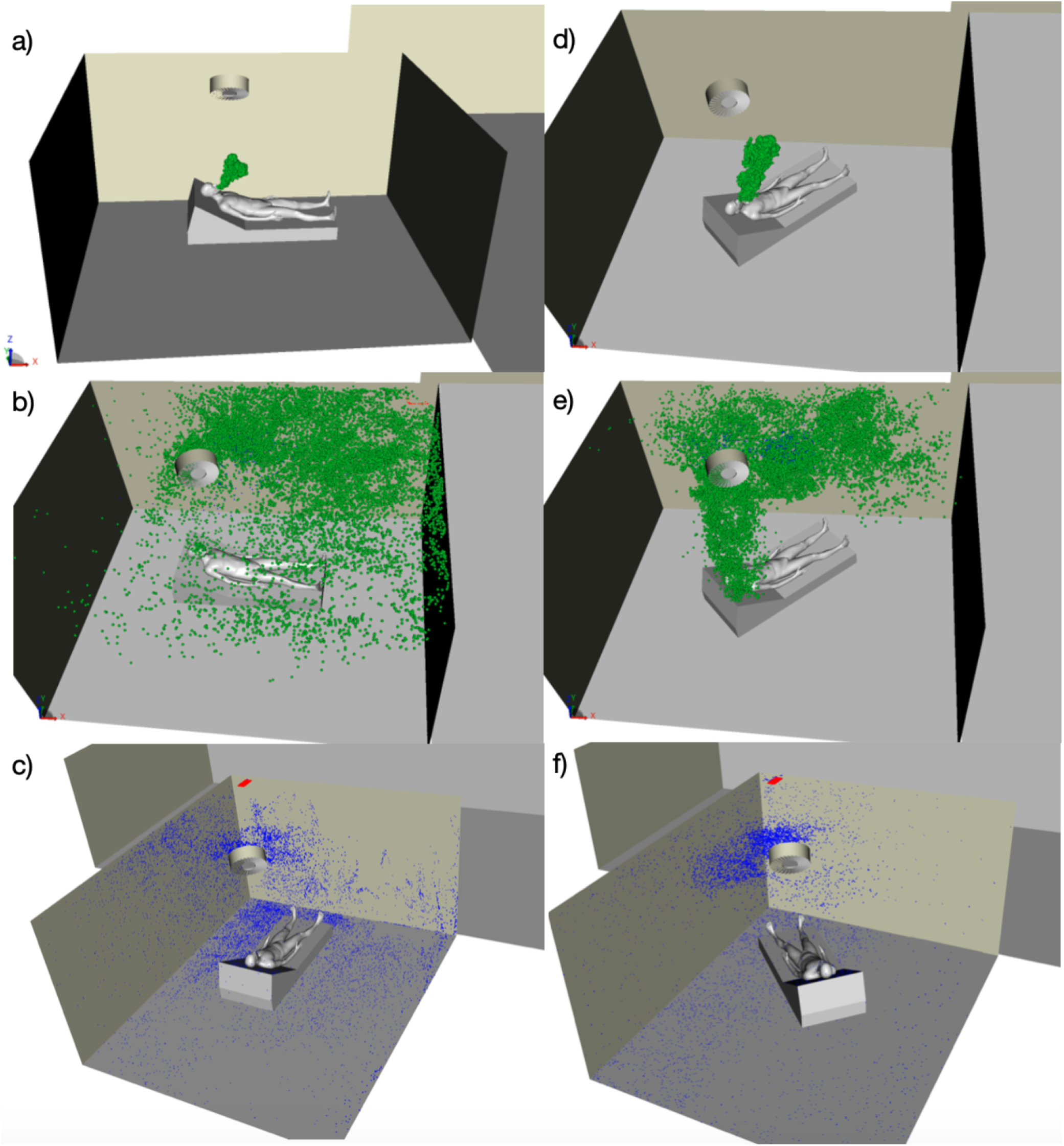
3D visualization of the numerically modeled airborne particles emitted by a patient coughing in an ICU room under negative pressure for the baseline configuration (left) and the 45° configuration (right): (a) & (d) At t = 5s; (b) & (e) At t = 20s; (c) & (f) Particles that are deposited at t = 45s. Airborne particles are in green, while deposited particles are in blue. The ventilation’s grey cylindrical inlet is above the patient’s head at the center of the ceiling and its red rectangular outlet is in the far left corner. Turning the bed by 45° towards the outlet helps reduce the dispersion of aerosols within the room, thus improving their extraction and concentrating deposits in a more reduced zone.

Based on these conclusions, simple mitigating scenarii were explored to reduce both deposit and airborne risks of viral transmission. The simplest method relies on changing the orientation of the patient’s bed from its baseline configuration to a 45° configuration aligned with the ventilation outlet. Similarly to the baseline configuration, the exhaled particles tend to go upwards initially (Fig. 4d). However, these airborne particles concentrate later on predominantly in the zone where the air extraction is located (Fig. 4e). It also avoids dispersing aerosols throughout the whole room. The impact points presented in Fig. 4f show a concentration of deposits on the left side of the room (where the extraction is positioned), mostly on the ceiling. The door area, which is on the right side of the room, shows a significant improvement when altering the bed position, with less deposits on the floor and on the door. Extra cleaning measures should be taken for surfaces presenting these higher concentration of deposit, mainly below the ventilation outlet. Lastly, the simulation results for the two additional configurations with an additional mobile air treatment unit are presented in the Supplementary information for the Plasmair behind the bed (Supplementary Fig. S12 and S13) and for the Plasmair in front of the bed (Supplementary Fig. S14 and S15).

In order to conclude on the best configuration choice, a quantitative analysis of the relative ratios between particles that remain airborne, those that are deposited on surfaces and those that are extracted through the ventilation is performed and results are presented in Fig. 5 for the four different configurations described previously. Fig. 5a indicates that by changing the orientation of the patient, over 30% more particles are extracted by the ventilation system as compared to the baseline configuration after 45s of time simulated. Extraction of aerosols can also be improved by 40% after 45s when adding a Plasmair in an optimized position (Fig. 1b). However, a wrong positioning of the Plasmair (Fig. 1d) can jeopardize its efficacy and lead to 65% fewer particles being extracted after 45s. Furthermore, Fig. 5b shows that the 45° bed orientation configuration does not reduce the number of particles that are deposited on surfaces throughout the room after 45 s as compared to the baseline configuration. However, adding a Plasmair behind the bed does reduce that amount of deposited particles by 25% after 45s. Overall, simple solutions can significantly reduce the amount of airborne particles emitted by a contagious patient within an ICU room, but if incorrectly implemented, can actually result in worsening the risk of contamination.

**Figure 5.**
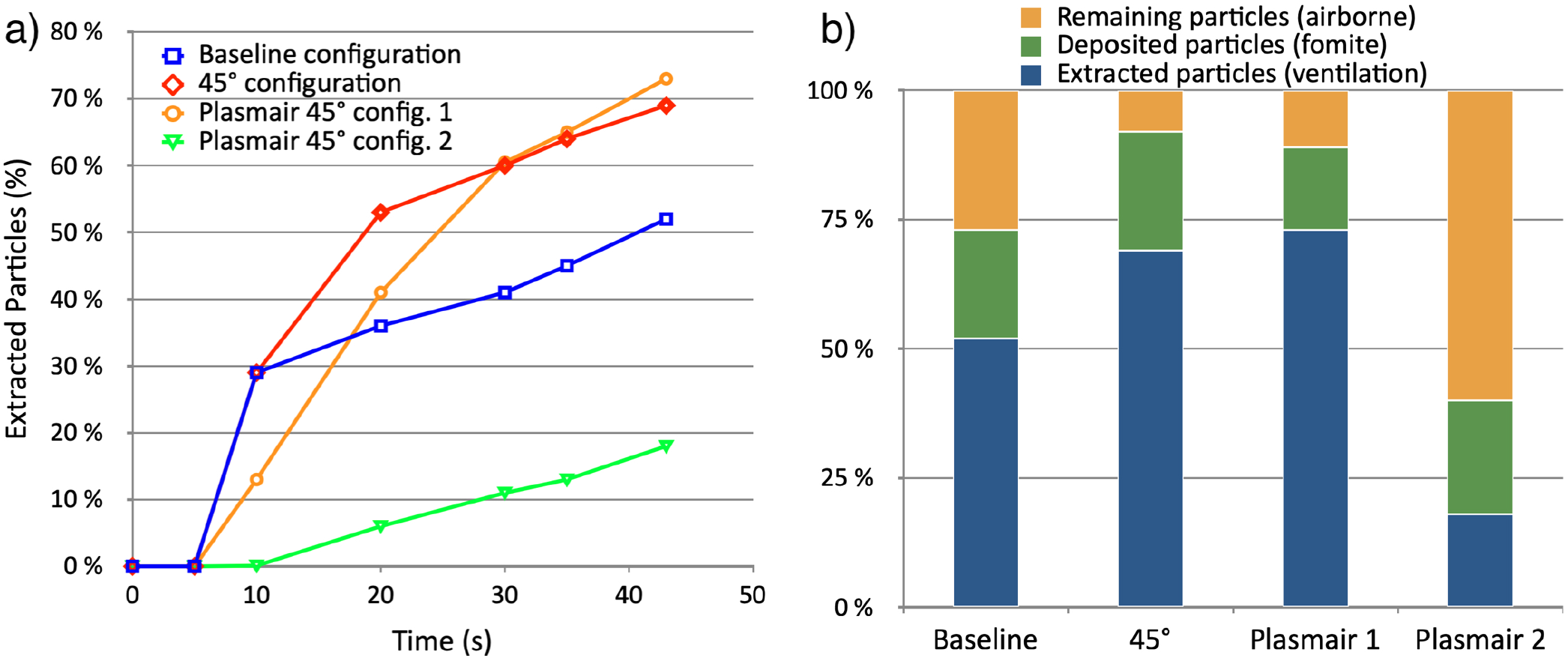
Influence of the room configuration on the extraction and deposition of aerosols. Graph (a) represents the evolution of the percentage of particles extracted through the ventilation system over time. The extraction rate is increased when changing the bed orientation and adding a Plasmair behind the bed. Graph (b) presents the ratios of extracted, deposited and airborne particles after 45s of physical time. The 45° bed position configuration is safer than the baseline one, as more particles are extracted by the ventilation (+32%). Adding a Plasmair in an optimal position can further improve the extraction of particles (+40%) and reduce the number of deposited particles (−25%) as compared to the baseline scenario. However an inadequate positioning can result in a consequent loss in extraction efficiency (−68%) after 45s.

## Discussion

Two recent studies have shown that COVID-19 patients can produce aerosols laden with viable SARS-CoV-2, even though no aerosol-generating procedure was in place^50^, and that these can travel long distances in hospital wards through ventilation^51^. This confirms the fact that the virus can be potentially transmitted via aerosols and that this must be taken into account when assessing preventive measures. A similar approach of CFD modelization of airflows was recently used to analyze the risk of infection between passengers in a car, with passive scalars modelling infectious pathogens^33^. It has yielded interesting results: opening all windows enables lowering the risk of transmission of airborne pathogens, demonstrating how crucial the study of airflows can be. Mitigating risks associated with aerosol propagation and potential transmission of pathogens to the medical personnel in hospitals is thus very relevant in the context of a pandemic such as the COVID-19 one. Different approaches to tackle that risk have been proposed: active measures (wearing Personal Protective Equipment and social distancing) as well as passive measures (air disinfection through Ultraviolet C lighting)^35^. But these are not always applicable in a medical context, especially in ICUs. It can thus be useful to use CFD to maximize the extraction of aerosolized pathogens by leveraging the rooms’ intrinsic airflows induced by HVAC.

Regarding the schlieren experiments, the volumetric flow rate presented in the supplementary documents is very close to the one chosen to model a cough described by Gupta, Lin & Chen^52^. The maximum velocities of exhaled air recorded are slightly lower than those found in the literature, where they can reach as much as 10 m/s^6,47^. Furthermore, the importance of the placement of the source of aerosols as well as the HVAC system within a ventilated room was mentioned in the literature^29–32,34^ and seems to be confirmed in our simulation. The size of particles also seems to have an impact on the extraction levels of aerosols through the HVAC^53^, something we did not observe in our study.

In order to optimize the model, it would be interesting to conduct precise particle sizing spectrometry on volunteers wearing HFNC and CPAP in order to confirm the approximations made. It is important to note that the schlieren technique yields information on the movement of exhaled air but not on the aerosols produced by a patient. The literature on the topic gives many different value ranges and no study has been made on oxygenated volunteers.

Regarding the schlieren technique, its main advantage is that it does not require the use of smoke and mannequins and can be performed on human volunteers, thus increasing reliability. However, the PIV analysis has some accuracy limitations when the contrast is not optimal^54^. Furthermore, only three young volunteers were used in the study, which does not represent the large variability in terms of manners of breathing and coughing.

A first limitation to the numerical model is the small time interval that was simulated. Simplifying the model would help increase the duration of the simulation and yield more complete results. However, many simplifications have already been made: evaporation is not taken into account in the model, the bed is the only piece of furniture modeled, and the LBM method does not yield a full picture of the 3-dimensional behavior of each particle. Due to the limited test data available on the flow field behavior inside the room, only the difference of pressure between the corridor and the room can be compared. The simulation provides a delta of 14 Pa when the test provides 15 Pa. To go deeper in the validation, air velocity measurements and smoke visualization can be conducted. Furthermore, the results presented here are adequate for the one room taken as an example. The same approach would have to be reused for each and every room having to be analyzed for risk mitigation.

A second limit to the model is that it assumes that every particle emitted by the patient contains virions. The virus has an elliptical shape, with a diameter ranging between 60 and 140 nm^18,55^. It can thus be present in micro-metric droplets, as long as their diameter is larger than 150 nm. However each droplet doesn’t necessarily contain a virion. In order to evaluate the probability of a nucleus being positive (ie: contain at least one virion and cause an infection according to the Independent Action Hypothesis (IAH)), it would be interesting to adapt the numerical model using a simple probabilistic volumetric model defined by a binomial law with the number of virions and droplet volume as parameters^48^. A 1 *µ*m droplet nucleus has a 0.01% chance of being positive, by assuming that the ratio between the diameter of a droplet before dehydration and the droplet’s nucleus is four^45^. It would be interesting to check if those estimations are correct by conducting in-situ experiments (Scanning Electron Microscopy, cascade impactor…). Finally, the assessment of the exposure time and/or the minimum viral load required to become infected would be relevant, as the IAH might be an oversimplification. The risk of infection could be found by using a dose-response equation, along with the total amount of virus present in the air and the exposure time^56^.

Furthermore, in order to confirm the numerical model’s validity, it would be interesting to collect air and surface samples in different areas of the room of an infected patient, as done to some extent by Santarpia et al.^26^ and Liu et al.^36^. A real time Reverse Transcriptase Polymerase Chain Reaction (rRT-PCR) analysis of these samples would yield results that could confirm the potential danger of aerosol propagation and corroborate the proposed mitigation strategies.

## Conclusions

It is fundamental for intensive care specialists to prevent as much as possible the cross transmission of SARS-CoV-2 to other patients and health care workers. For severe patients with ARF, wearing a face mask is difficult if not impossible. Therefore, the risk of airborne transmission should not be minimized. Our study provides arguments for an extreme prudence in using negative pressure in ICU rooms and gives important messages on the importance of adequately positioning the patients and mobile air treatment units within these rooms. It also highlights key elements that need to be taken into account when designing single hospital rooms in order to optimize the extraction of airborne pathogens. In order to do so, our study provides a CFD method that can be easily replicated and applied to any existing room as well as to design new room configurations for future hospitals.

Secondly, the schlieren technique is adequate in order to visualize qualitatively and quantitatively the airflows produced by different types of respiratory events depending on the oxygenation technique: unlike Continuous Positive Airway Pressure masks, High Flow Nasal Canulae notably increase the flow rate of air exhaled by the patient, potentially increasing the shedding of infectious airborne particles. These results were used to calibrate the numerical model to describe the fluid dynamics and aerosol propagation within the room. The comparison between the model and the schlieren images indicates a good correlation.

Thirdly, the use of numerical modeling can prove critical in order to analyse the movements of particles exhaled by an infected patient within an ICU room, test different mitigating scenarios, and reduce risk for medical personnel entering the room. The application of a code based on the Lattice Boltzmann Method to this simulation enables following precisely the movements of large numbers of airborne particles along airflows in surroundings that can be modeled simply and interchangeably. Simple measures, when applied correctly, can reduce risk of transmission by increasing the extraction rate of airborne particles through the ventilation system and by reducing the amount of particles deposited on surfaces. Numerical results for a typical ICU room indicate that adequate bed orientation can improve by over 30% the number of particles extracted after 45 s. The positioning of an additional air treatment unit can help that number reach 40%, although it can cause a deterioration of 65% if inadequately placed. However, each room having its own parameters, the use of such a method implies having to proceed with a case by case approach.

## Methods

In order to model the air flows associated with different respiratory events, we chose to visualize and characterize them through the schlieren method applied to biomedical imaging as described by Tang et al.^25^. The volunteer is placed between a light source and a spherical mirror. He exhales gas whose temperature is higher than the surrounding room’s temperature. Changes in gas temperature lead to changes in air density, which in turn alter the refractive index. A light beam that goes through a volume of air with varying temperature along its path is thus refracted and bent in a way that can be recorded as a grey-scale image when subtracting the unrefracted light by using a spatial filter, here a razor blade^25^. The first phase of this method is the observation of a cough or breath with a high speed camera (2000 fps). In a second phase, the images were analyzed by pairs using the digital particle image velocimetry algorithm ‘PIVlab’ in order to determine the evolution of flow rates over time of the exhaled air, using the turbulent patterns as tracers^54^. Results are shown on a map of vectors.

The study was approved by the CPP Ile de France XI ethics committee (Comité de Protection des Personnes, N° IDRCB: 2020-001457-43, Ref. CPP: 20037-25957) as a nested study of the COVIDICUS randomized controlled trial (APHP 200388-COVIDICUS). All experiments were performed in accordance with relevant guidelines and regulations. Informed consent was obtained from patients or surrogates of Bichat hospital that will serve for modeling aerosol propagation. Measurements of the flow rate values were done on three healthy subjects in the 20-30 age range, breathing or coughing under three different scenarios: without HFNC, with HFNC at 30 L/min of added oxygen, and with HFNC at 60 L/min. The model used was Fisher & Paykel Healthcare’s Optiflow™ Nasal High Flow Therapy delivered by AIRVO™ 2. Measurements were also done with CPAP at 15 L/min and 30 L/min of added oxygen. The model used was Vygon’s CPAP Boussignac and anti-microbial filters were fitted at the end of the system. It is assumed that all virus-laden particles are stopped by this filter; the airflows emanating from their exit were not studied.

The numerical results presented in this study were generated using the commercial Lattice Boltzmann-based Method (LBM) code, PowerFLOW®^57,58^. Instead of solving the Navier Stokes equations, this code solves a form of the Boltzmann equation in a discretized velocity space. The Navier Stokes equations can be derived from the Boltzmann equation using the Chapman-Enskog Expansion^59^. The turbulence scheme used in this study is the Very Large Eddy Simulation (VLES) approach^58^. PowerFLOW® has an integrated Lagrangian particle simulator. This simulator includes models for splash breakup and re-entrainment. The splash model is based on the work of Mundo, Sommerfeld & Tropea^60^ and O’Rourke, and Amsden^61^, while the breakup model is derived from the TAB model^62^. Lagrangian particle tracking in CFD has been the subject of intense research for decades and are well summarized in several works^63–67^. In the current study, the trajectory of the Lagrangian particles is predicted using Eq.(1):

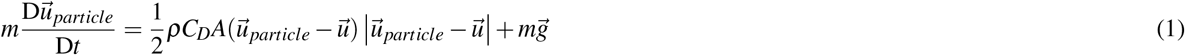

where m is the particle mass, 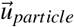 is the particle velocity, *C*_*D*_ is the particle drag coefficient (dependent on the velocity), and A is the particle’s cross sectional area. The drag force is also accounted for as a reactionary force on the fluid, bi-directionally coupling the particle’s momentum with the fluid’s momentum. This coupling allows the trajectory of millions of particles to be accurately tracked. The drag coefficient is obtained using the classical law coupling the drag coefficient to the local Reynolds number^63,64^ as shown in Eq.(2):

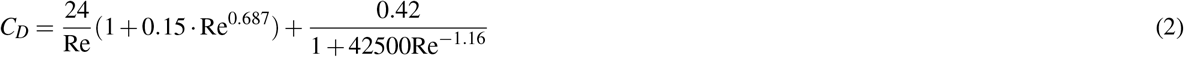

using the local Reynolds number Re defined by Eq.(3):

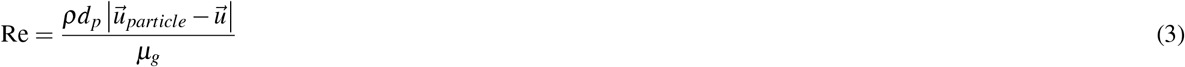

where *d*_*p*_ is the particle size, while *ρ*_*g*_ and *µ*_*g*_ are the air density and dynamical viscosity respectively. This relation for the drag coefficient accounts well for the different drag regimes, namely Stokes drag for low relative velocity (linear relation for small Reynolds number) and Newton law (drag proportional to the square of the relative velocity) at large Reynolds number. Furthermore, one can define the Stokes number of the particles as the ratio between the particle inertia and the air viscous drag as presented in Eq.(4):

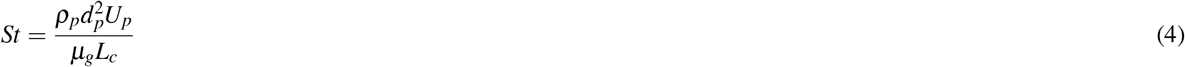

where *ρ*_*p*_ is the particle density, *U*_*p*_ its typical velocity and *L*_*c*_ the typical scale of the flow (of the order of the mouth size here). For our simulations, the Stokes number is always smaller than 0.005 indicating that the particle inertia remains always small as compared to the Stokes drag and more precisely that the particles follow mostly the air flow, as described by the model. This range is consistent with other published results for breathing/coughing events^68^.

The information required to set up the cough simulations shown in this paper was collected from several sources. The cough is a double cough with the flow rate variation over time taken from Gupta, Lin & Chen^52^ (see Supplementary Fig. S1 and S2 online). The mouth opening size was approximately 5 cm^2^, which is the average mouth opening size recorded for males^52^. In order to be able to predict the right flow exiting the mouth, it is key to get a correct level of flow velocity, turbulence and jet angle while the air is developing after the mouth. A high fidelity human model was used including a 3D model of the throat, teeth, tongue and mouth as shown in the supplementary documents (Supplementary Fig. S10 and S11).

To ensure a good accuracy for the simulation, some parameters are important to describe. The maximum velocity of the air exiting the mouth is around 10 m/s. This corresponds to a low Mach number, which is within the validity range of the Lattice Boltzmann solver. The mesh refinement is key to represent properly the flow field and the finest structures. The finest cell size used is 1 mm, which guaranties a section of at least 10 cells in the throat of the virtual mannequin. Different spheres of resolution are set around the mouth in order to predict properly the development of the flow exiting the mouth: 2 mm, 4 mm and so on… (Supplementary Fig. S12). Moreover, a wall model is used throughout the simulation. The Y+ map around the mannequin highlights a very good resolution around the mannequin and inside the room with Y+ values under 20 and as low as under 5 in the mouth (Supplementary Fig. S13). In terms of mesh statistics, the simulation is split between volume cells (called Voxels) and surface cells (Surfels). The simulation comprises approximately 52 million Voxels and 2,7 million Surfels. This level of detail, added to a high fidelity LBM solver, yields a satisfactory accuracy level of the predicted flow field in a reasonable computational time.

The room used for the simulation is under negative pressure, with a measured pressure difference of 15 Pa between it and the corridor. The ventilation system is finely reproduced based on technical specifications. It presents an air mass flow inlet at the center of the room’s ceiling, which injects 788 m^3^/h just above the patient’s bed head. Additionally, an outlet extracting 1182 m^3^/h is located in the left corner in front of the patient. The pressure difference was not input in the model as it is a resultant of the simulation and is maintained by a combination of different parameters. Firstly, 50% more air is extracted from the room than is blown in through the ventilation. Furthermore, an ambient atmospheric pressure was set in the corridor behind the room’s door. Finally, the remaining air that has to be extracted by the outlet in order to stabilize the pressure inside the simulation model is injected from the corridor, through the 10 mm gap that exists between the door and the floor. Since the corridor pressure will remain at 101325 Pa, the pressure difference in the ICU room is a direct result from the simulation. The simulation provided a pressure difference of 14 Pa, which was very close to the in-situ measurement. To properly assess both the pressure difference and the flow field inside the ICU room, a fine mesh strategy was used: a 1 mm resolution was used for cells around the HVAC inlet, and a 2 mm resolution was used around the gap under the door. Since a high velocity jet was expected under it, additional resolutions were set to allow a good development of flow structures. Finally the maximum resolution used in the rest of the room was 2,4 cm. After an initial phase where the cough plays the major role in the dispersion and direction of the exhaled particles, the ventilation system and the flow field around the patient are key to understanding how the particles travel. Finally, the additional air treatment unit that was used in parts of the simulation is modeled on the characteristics of the PLASMAIR® Guardian: the air enters from below at a flow of 1000 m^3^/h, and is then disinfected before being re-injected in the room through its top.

In order to correctly model the movement of aerosols within the room, we took values based on the available literature to determine the appropriate numbers for size distribution and number of particles emitted during respiratory events. We chose not to model the larger-sized particles which do not remain airborne long enough. We assumed that the size distribution of aerosols produced when breathing or coughing with HFNC is similar to the size distribution of those produced when breathing normally, reduced to their droplet nuclei. The particle size distribution used in the model is a Normal Distribution fitted to the diameter data of Zayas et al.^41^ (see Supplementary Fig. S3 online), which is centered around 0,3 *µ*m. Furthermore, given the dispersion in values for the average number of exhaled particles per litre, we chose to model 1 × 10^5^ exhaled particles per second when considering a patient breathing with HFNC. Lastly, the initial particle velocity distribution was not seen to have a large impact on the results of the simulations due to the small Stokes numbers of the particles and the large amount of mixing which occurs before exiting the mouth.

## Data Availability

All raw data used for analysis are available and have been deposited at Zenodo.

https://doi.org/10.5281/zenodo.4056819

## Acknowledgements

We gratefully thank the staff of the Intensive Care Unit at Bichat Hospital, particularly Delphine Saint-Leandre, for their time, advice and lending of medical equipment. We would also like to acknowledge the technical help from Dassault Systèmes: advice, software and computing time, as well as proof-reading from Gregory Laskowski. Finally, we thank the LadHyx and the experimental labs of the department of Mechanics at Ecole polytechnique for their support and equipment for the optical experiments, in particular William Gilbert.

## Author contributions statement

C.D., C.J., L.B. and J.-F.T. designed the research and provided technical expertise. C.C., M.L., B.D., E.V., J.J. conducted the experiments and simulations. C.C., B.D. and E.V. analyzed the results. All authors reviewed the manuscript.

## Additional information

### Data availability

Supplementary movies are available at https://doi.org/10.5281/zenodo.4056819.

### Competing interests

Jean-François Timsit - For the submitted work: None to declare - Outside the submitted work: advisory board participations (Merck, Pfizer, Bayer, Nabriva, Paratek, BD); lectures (Merck, Pfizer, Biomerieux); grants to research unit (Merck, Pfizer, 3M).

## Notes

### Competing Interest Statement

Jean-Francois Timsit:
- For the submitted work: None to declare.
- Outside the submitted work: advisory board participations (Merck, Pfizer, Bayer, Nabriva, Paratek, BD); lectures (Merck, Pfizer, Biomerieux); grants to research unit (Merck, Pfizer, 3M).

### Funding Statement

No external funding was received.

### Author Declarations

IRB : Comite de Protection des Personnes (CCP Ile de France XI) IDRCB number : 2020-001457-43 Ref. CPP : 20037-25957 Ethical approval was given

## References

1. WHO. Modes of transmission of virus causing covid-19: implications for ipc precaution recommendations (2020).

2. WHO. Coronavirus disease (covid-2019) situation reports (2021).

3. Prather, K. A., Wang, C. C. & Schooley, R. T. Reducing transmission of sars-cov-2. Science eabc6197, DOI: 10.1126/science.abc6197 (2020).

4. Zhang, R., Li, Y., Zhang, A. L., Wang, Y. & Molina, M. J. Identifying airborne transmission as the dominant route for the spread of covid-19. Proc. Natl. Acad. Sci. 202009637, DOI: 10.1073/pnas.2009637117 (2020).

5. Tellier, R. Review of aerosol transmission of influenza a virus. Emerg. infectious diseases 12, 1657–1662, DOI: 10.3201/eid1211.060426 (2006).

6. Xie, X., Li, Y., Chwang, A. T. Y., Ho, P. L. & Seto, W. H. How far droplets can move in indoor environments – revisiting the wells evaporation–falling curve. Indoor air 17, 211–225, DOI: 10.1111/j.1600-0668.2007.00469.x (2007).

7. Morawska, L. & Cao, J. Airborne transmission of sars-cov-2: The world should face the reality, DOI: https://doi.org/10.1016/j.envint.2020.105730 (2020).

8. Bourouiba, L. Turbulent gas clouds and respiratory pathogen emissions: Potential implications for reducing transmission of covid-19. JAMA 323, 1837, DOI: 10.1001/jama.2020.4756 (2020).

9. van Doremalen, N. et al. Aerosol and surface stability of sars-cov-2 as compared with sars-cov-1. N Engl J Med 382, 1564–1567, DOI: 10.1056/NEJMc2004973 (2020).

10. Li, Y. et al. Evidence for probable aerosol transmission of sars-cov-2 in a poorly ventilated restaurant. medRxiv 2020.04.16.20067728, DOI: 10.1101/2020.04.16.20067728 (2020).

11. Liu, Y. et al. Aerodynamic analysis of sars-cov-2 in two wuhan hospitals. Nature DOI: 10.1038/s41586-020-2271-3 (2020).

12. Kluytmans-van den Bergh, M. F. Q. et al. Prevalence and clinical presentation of health care workers with symptoms of coronavirus disease 2019 in 2 dutch hospitals during an early phase of the pandemic. JAMA network open 3, e209673, DOI: 10.1001/jamanetworkopen.2020.9673 (2020).

13. Chou, R. et al. Epidemiology of and risk factors for coronavirus infection in health care workers. Annals Intern. Medicine 173, 120–136, DOI: 10.7326/M20-1632 (2020).

14. Çelebi, G. et al. Specific risk factors for sars-cov-2 transmission among health care workers in a university hospital. Am. J. Infect. Control. DOI: 10.1016/j.ajic.2020.07.039 (2020).

15. Galanis, P., Vraka, I., Fragkou, D., Bilali, A. & Kaitelidou, D. Seroprevalence of sars-cov-2 antibodies and associated factors in health care workers: a systematic review and meta-analysis. J. Hosp. Infect. DOI: https://doi.org/10.1016/j.jhin.2020.11.008 (2020).

16. Dimcheff, D. E. et al. Seroprevalence of severe acute respiratory syndrome coronavirus-2 (sars-cov-2) infection among veterans affairs healthcare system employees suggests higher risk of infection when exposed to sars-cov-2 outside the work environment. Infect. Control. Hosp. Epidemiol. 1–7, DOI: 10.1017/ice.2020.1220 (2020).

17. Steensels, D. et al. Hospital-wide sars-cov-2 antibody screening in 3056 staff in a tertiary center in belgium. JAMA 324, 195–197, DOI: 10.1001/jama.2020.11160 (2020).

18. Wiersinga, W. J., Rhodes, A., Cheng, A. C., Peacock, S. J. & Prescott, H. C. Pathophysiology, transmission, diagnosis, and treatment of coronavirus disease 2019 (covid-19). J. Am. Med. Assoc. DOI: 10.1001/jama.2020.12839 (2020).

19. Ng, K. et al. Covid-19 and the risk to health care workers: A case report. Annals Intern. Medicine 172, 766–767, DOI: 10.7326/L20-0175 (2020).

20. Auld, S. et al. Icu and ventilator mortality among critically ill adults with covid-19. medRxiv 2020.04.23.20076737, DOI: 10.1101/2020.04.23.20076737 (2020).

21. Tobin, M. J. Basing respiratory management of covid-19 on physiological principles. Am J Respir Crit Care Med 201, 1319–1320, DOI: 10.1164/rccm.202004-1076ED (2020).

22. Bellani, G. et al. Noninvasive ventilation of patients with acute respiratory distress syndrome. insights from the lung safe study. Am J Respir Crit Care Med 195, 67–77, DOI: 10.1164/rccm.201606-1306OC (2017).

23. Bouadma, L., Lescure, F.-X., Lucet, J.-C., Yazdanpanah, Y. & Timsit, J.-F. Severe sars-cov-2 infections: practical considerations and management strategy for intensivists. Intensive Care Medicine 46, 579–582, DOI: 10.1007/s00134-020-05967-x (2020).

24. Winck, J. C. & Ambrosino, N. Covid-19 pandemic and non invasive respiratory management: Every goliath needs a david. an evidence based evaluation of problems. Pulmonology DOI: 10.1016/j.pulmoe.2020.04.013 (2020).

25. Tang, J. W., Liebner, T. J., Craven, B. A. & Settles, G. S. A schlieren optical study of the human cough with and without wearing masks for aerosol infection control. J. The Royal Soc. Interface 6, S727–S736, DOI: 10.1098/rsif.2009.0295.focus (2009).

26. Santarpia, J. L. et al. Aerosol and surface transmission potential of sars-cov-2. medRxiv 2020.03.23.20039446, DOI: 10.1101/2020.03.23.20039446 (2020).

27. Wang, K., Zhao, W., Li, J., Shu, W. & Duan, J. The experience of high-flow nasal cannula in hospitalized patients with 2019 novel coronavirus-infected pneumonia in two hospitals of chongqing, china. Annals Intensive Care 10, 37, DOI: 10.1186/s13613-020-00653-z (2020).

28. Lemiale, V. et al. Effect of noninvasive ventilation vs oxygen therapy on mortality among immunocompromised patients with acute respiratory failure: A randomized clinical trial. J. Am. Med. Assoc. 314, 1711–1719, DOI: 10.1001/jama.2015.12402 (2015).

29. Bhagat, R. K., Wykes, M. S. D., Dalziel, S. B. & Linden, P. F. Effects of ventilation on the indoor spread of covid-19. J. Fluid Mech. 903, F1, DOI: 10.1017/jfm.2020.720 (2020).

30. Kao, P. H. & Yang, R. J. Virus diffusion in isolation rooms. J. Hosp. Infect. 62, 338–345, DOI: https://doi.org/10.1016/j.jhin.2005.07.019 (2006).

31. Mao, S. & Celik, I. B. Modeling of indoor airflow and dispersion of aerosols using immersed boundary and random flow generation methods. Comput. & Fluids 39, 1275–1283, DOI: https://doi.org/10.1016/j.compfluid.2010.03.010 (2010).

32. Redrow, J., Mao, S., Celik, I., Posada, J. A. & gang Feng, Z. Modeling the evaporation and dispersion of airborne sputum droplets expelled from a human cough, DOI: https://doi.org/10.1016/j.buildenv.2011.04.011 (2011).

33. Mathai, V., Das, A., Bailey, J. A. & Breuer, K. Airflows inside passenger cars and implications for airborne disease transmission. Sci Adv 7, eabe0166, DOI: 10.1126/sciadv.abe0166 (2021).

34. Dbouk, T. & Drikakis, D. On coughing and airborne droplet transmission to humans. Phys. Fluids 32, 053310, DOI: 10.1063/5.0011960 (2020).

35. Buchan, A. G., Yang, L. & Atkinson, K. D. Predicting airborne coronavirus inactivation by far-uvc in populated rooms using a high-fidelity coupled radiation-cfd model. Sci. Reports 10, 19659, DOI: 10.1038/s41598-020-76597-y (2020).

36. Liu, H., He, S., Shen, L. & Hong, J. Simulation-based study of covid-19 outbreak associated with air-conditioning in a restaurant. Phys. Fluids 33, 023301, DOI: 10.1063/5.0040188 (2021). Doi: 10.1063/5.0040188; 30.

37. Chong, K. L. et al. Extended lifetime of respiratory droplets in a turbulent vapor puff and its implications on airborne disease transmission. Phys. Rev. Lett. 126, 034502, DOI: 10.1103/PhysRevLett.126.034502 (2021). ID: 10.1103/Phys- RevLett.126.034502; J1: PRL.

38. Lindsley, W. et al. Quantity and size distribution of cough-generated aerosol particles produced by influenza patients during and after illness. J. occupational environmental hygiene 9, 443–9, DOI: 10.1080/15459624.2012.684582 (2012).

39. Morawska, L. et al. Size distribution and sites of origin of droplets expelled from the human respiratory tract during expiratory activities, DOI: https://doi.org/10.1016/j.jaerosci.2008.11.002 (2009).

40. Lee, J. et al. Quantity, size distribution, and characteristics of cough-generated aerosol produced by patients with an upper respiratory tract infection. Aerosol Air Qual. Res. 19, 840–853, DOI: 10.4209/aaqr.2018.01.0031 (2019).

41. Zayas, G. et al. Cough aerosol in healthy participants: fundamental knowledge to optimize droplet-spread infectious respiratory disease management. BMC Pulm. Medicine 12, 11, DOI: 10.1186/1471-2466-12-11 (2012).

42. Balachandar, S., Zaleski, S., Soldati, A., Ahmadi, G. & Bourouiba, L. Host-to-host airborne transmission as a multiphase flow problem for science-based social distance guidelines. Int. J. Multiph. Flow 132, 103439, DOI: https://doi.org/10.1016/j.ijmultiphaseflow.2020.103439 (2020).

43. Han, Z. Y., Weng, W. G. & Huang, Q. Y. Characterizations of particle size distribution of the droplets exhaled by sneeze. J. Royal Soc. Interface 10, DOI: https://doi.org/10.1098/rsif.2013.0560 (2013).

44. Papineni, R. & Rosenthal, F. The size distribution of droplets in the exhaled breath of healthy human subjects. J. Aerosol Medicine 10, 105–116, DOI: 10.1089/jam.1997.10.105 (1997).

45. Liu, L., Wei, J., Li, Y. & Ooi, A. Evaporation and dispersion of respiratory droplets from coughing. Indoor air 27, 179–190, DOI: 10.1111/ina.12297 (2017).

46. Morawska, L. Droplet fate in indoor environments, or can we prevent the spread of infection? Indoor air 16, 335–347, DOI: 10.1111/j.1600-0668.2006.00432.x (2006).

47. Mittal, R., Ni, R. & Seo, J.-H. The flow physics of covid-19. J. Fluid Mech. 894, F2, DOI: 10.1017/jfm.2020.330 (2020).

48. Stadnytskyi, V., Bax, C. E., Bax, A. & Anfinrud, P. The airborne lifetime of small speech droplets and their potential importance in sars-cov-2 transmission. Proc Natl Acad Sci USA 117, 11875, DOI: 10.1073/pnas.2006874117 (2020).

49. Bourouiba, L. The fluid dynamics of disease transmission. Annu. Rev. Fluid Mech. 53, 473–508, DOI: 10.1146/annurev-fluid-060220-113712 (2021). Doi: 10.1146/annurev-fluid-060220-113712; 30.

50. Lednicky, J. A. et al. Viable sars-cov-2 in the air of a hospital room with covid-19 patients. medRxiv 2020.08.03.20167395, DOI: 10.1101/2020.08.03.20167395 (2020).

51. Nissen, K. et al. Long-distance airborne dispersal of sars-cov-2 in covid-19 wards. Sci. Reports 10, 19589, DOI: 10.1038/s41598-020-76442-2 (2020).

52. Gupta, J. K., Lin, C. H. & Chen, Q. Flow dynamics and characterization of a cough. Indoor air 19, 517–525, DOI: 10.1111/j.1600-0668.2009.00619.x (2009).

53. Grosskopf, K. R. & Herstein, K. R. The aerodynamic behavior of respiratory aerosols within a general patient room. Null 18, 709–722, DOI: 10.1080/10789669.2011.587586 (2012).

54. Thielicke, W. & Stamhuis, E. J. Pivlab – towards user-friendly, affordable and accurate digital particle image velocimetry in matlab. J. Open Res. Softw. 2, e30 (2014).

55. Cascella, M., Rajnik, M., Cuomo, A., Dulebohn, S. & Napoli, R. D. Evaluation and treatment coronavirus (covid-19). StatPearls (2020).

56. Adhikari, U. et al. A case study evaluating the risk of infection from middle eastern respiratory syndrome coronavirus (mers-cov) in a hospital setting through bioaerosols. Risk Analysis 39, 2608–2624, DOI: 10.1111/risa.13389 (2019).

57. Chen, S. & Doolen, G. D. Lattice boltzmann method for fluid flows. Annu. Rev. Fluid Mech. 30, 329–364, DOI: 10.1146/annurev.fluid.30.1.329 (1998).

58. Chen, H. et al. Extended boltzmann kinetic equation for turbulent flows. Science 301, 633–636, DOI: 10.1126/science.1085048 (2003).

59. Williams, M. R. Mathematical theory of transport processes in gasesj. h. ferziger and h. g. kaper, north holland, amsterdam (1972). pp. 579 (1973).

60. Mundo, C., Sommerfeld, M. & Tropea, C. Droplet-wall collisions: Experimental studies of the deformation and breakup process. Int. J. Multiph. Flow 21, 151–173, DOI: https://doi.org/10.1016/0301-9322(94)00069-V (1995).

61. O’Rourke, P. J. & Amsden, A. A. A spray/wall interaction submodel for the kiva-3 wall film model, DOI: 10.4271/2000-01-0271 (2000).

62. O’Rourke, P. J. & Amsden, A. A. The tab method for numerical calculation of spray droplet breakup.. (1987).

63. Clift, R., Grace, J. & Weber, M. Bubbles, Drops and Particles (Academic Press, New York, 1978).

64. Ho, C. A. & Sommerfeld, M. Modelling of micro-particle agglomeration in turbulent flows. Chem. Eng. Sci. 57, 3073–3084 (2002).

65. Toschi, F. & Bodenschatz, E. Lagrangian properties of particles in turbulence. Annu. Rev. Fluid Mech. 41, 375–404, DOI: 10.1146/annurev.fluid.010908.165210 (2009). Doi: 10.1146/annurev.fluid.010908.165210; 31.

66. Mathai, V., Lohse, D. & Sun, C. Bubbly and buoyant particle–laden turbulent flows. Annu. Rev. Condens. Matter Phys. 11, 529–559, DOI: 10.1146/annurev-conmatphys-031119-050637 (2020). Doi: 10.1146/annurev-conmatphys-031119-050637; 31.

67. Balachandar, S. & Eaton, J. K. Turbulent dispersed multiphase flow. Annu. Rev. Fluid Mech. 42, 111–133, DOI: 10.1146/annurev.fluid.010908.165243 (2010). Doi: 10.1146/annurev.fluid.010908.165243; 31.

68. Rosti, M. E., Cavaiola, M., Olivieri, S., Seminara, A. & Mazzino, A. Turbulence role in the fate of virus-containing droplets in violent expiratory events. Phys. Rev. Res. 3, 013091, DOI: 10.1103/PhysRevResearch.3.013091 (2021).

