## Supplementary Information for "Modeling of aerosol transmission of airborne pathogens in ICU rooms of COVID-19 patients with acute respiratory failure" (from another version of this preprint: version 1, 2020-12-14)

**This PDF file includes:**

Figures S1 to S17
Legends for Videos S1 to S3
SI References

**Other supplementary materials for this manuscript include the following:**

Movies S1 to S3

**Figures**

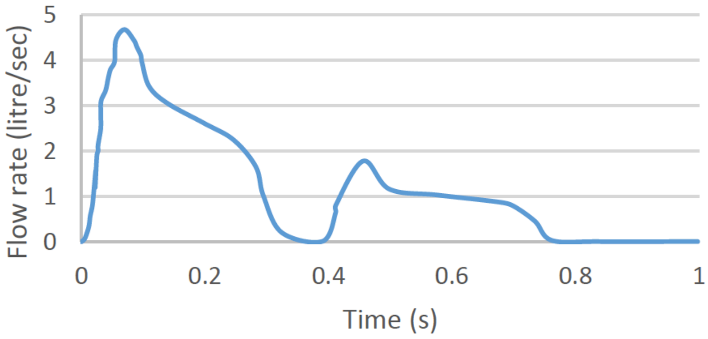


**Supplementary Fig. S1.** Cough volumetric flow rate used in the simulation (Gupta et al., 2009)

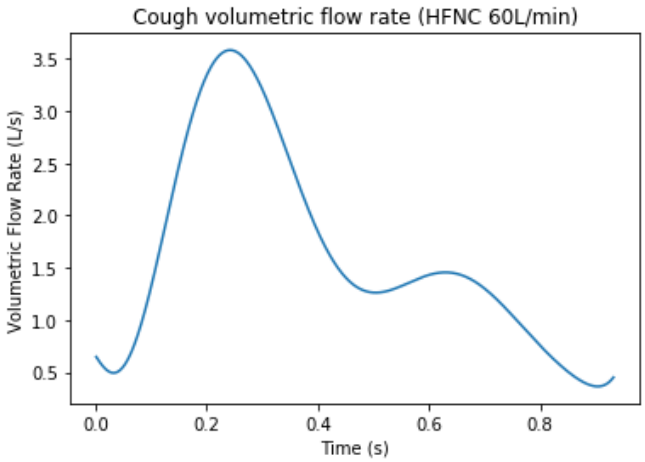


**Supplementary Fig. S2.** Quantitative result from Schlieren images of a volunteer’s cough with HFNC at 60L/min after polynomial interpolation. The cough volumetric flow rate is estimated through PIV analysis, extrapolation and integration of all video frames. It is similar to the one used in the model: identical shape with two peaks, comparable volume of exhaled air.

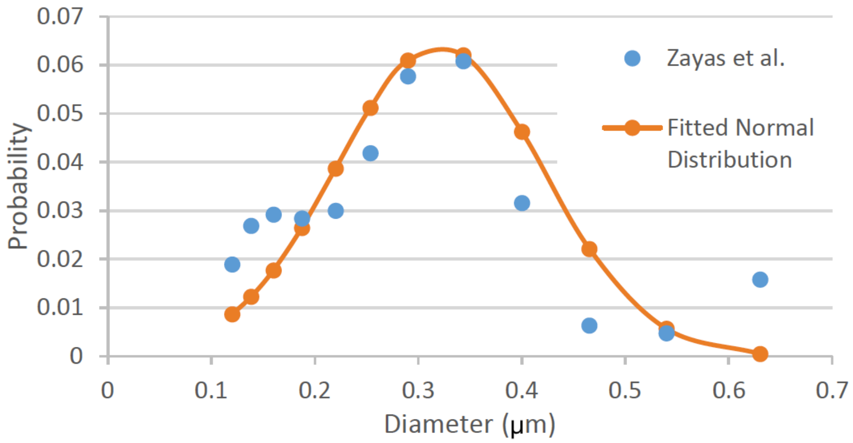


**Supplementary Fig. S3.** Particle size distribution used in the simulation (Zayas et al., 2012)

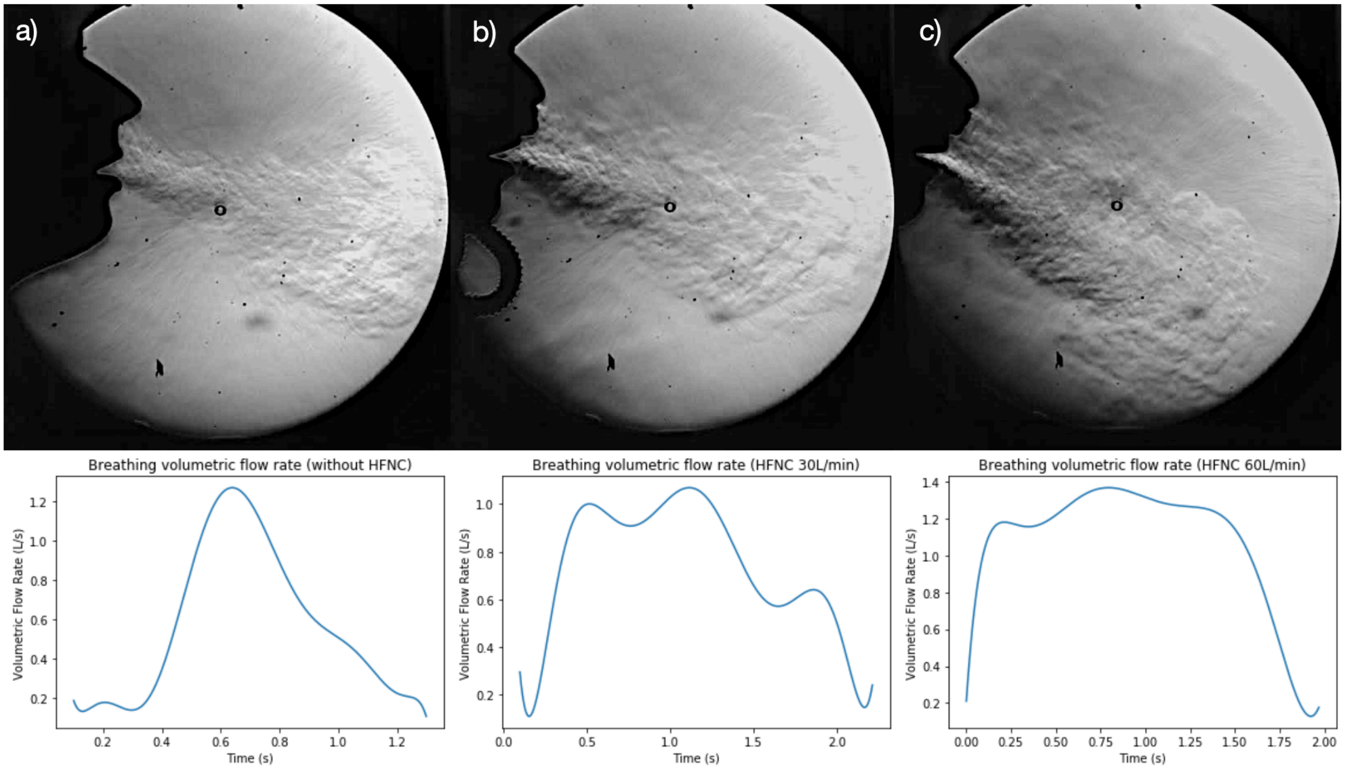


**Supplementary Figure S4.** Schlieren side view images and associated volumetric flow rate estimations of a volunteer breathing a) without HFNC; b) with HFNC at 30L/min; c) with HFNC at 60L/min. The higher the input volume of air through HFNC, the more air is exhaled at higher speeds and flow rates.

| Breathing type | Average flow rate (L/s) |
|---|---|
| **Normal** | 0,48 ± 0,07 |
| **HFNC 30 L/min** | 0,94 ± 0,16 |
| **HFNC 60 L/min** | 1,24 ± 0,05 |

**Supplementary Figure S5.** Average flow rate values obtained for different types of breathing after PIV analysis of the Schlieren videos of 3 volunteers

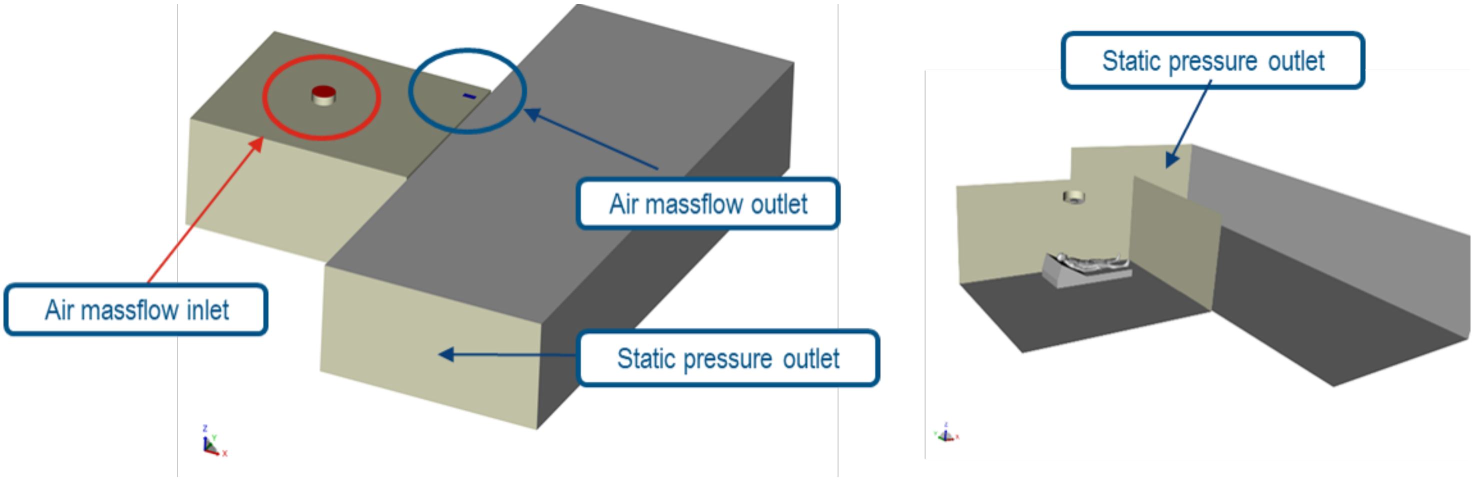


**Supplementary Fig. S6.** Boundary conditions of the simulation and ICU room geometry showing the position of the ventilation inlet and outlet

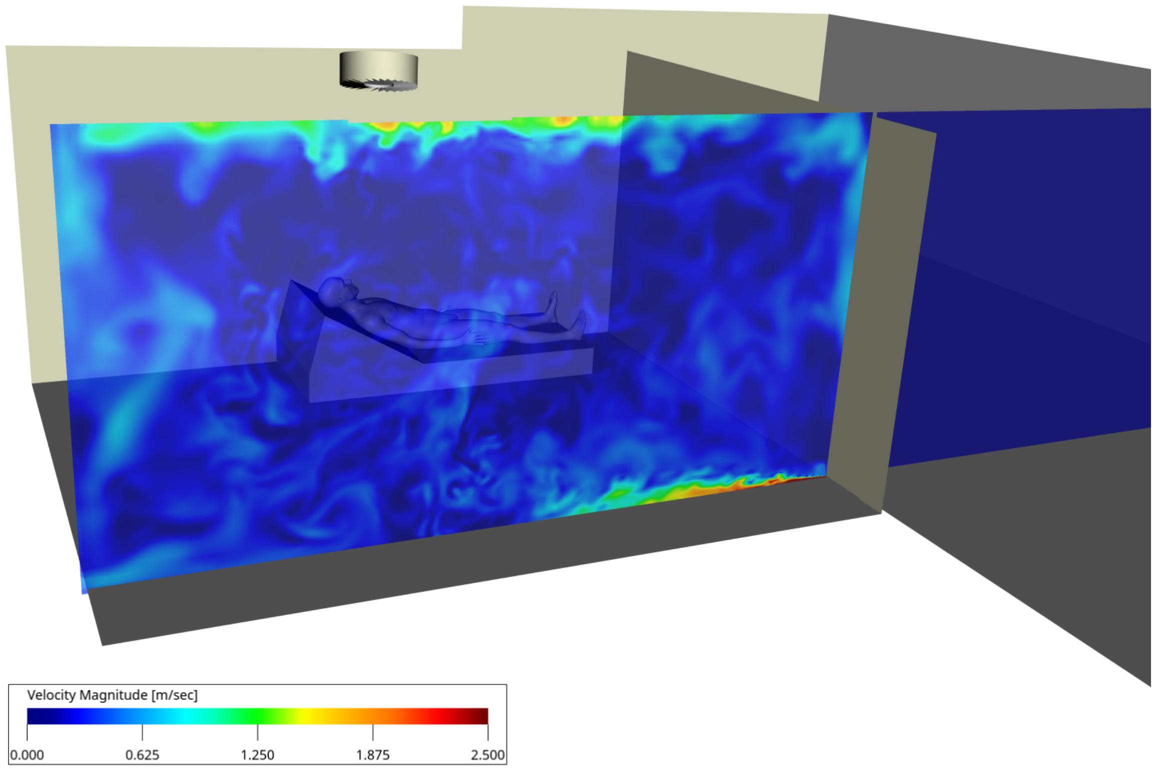


**Supplementary Fig. S7.** 3D visualization presenting the simulated air velocity magnitude at the door location and Y constant position.

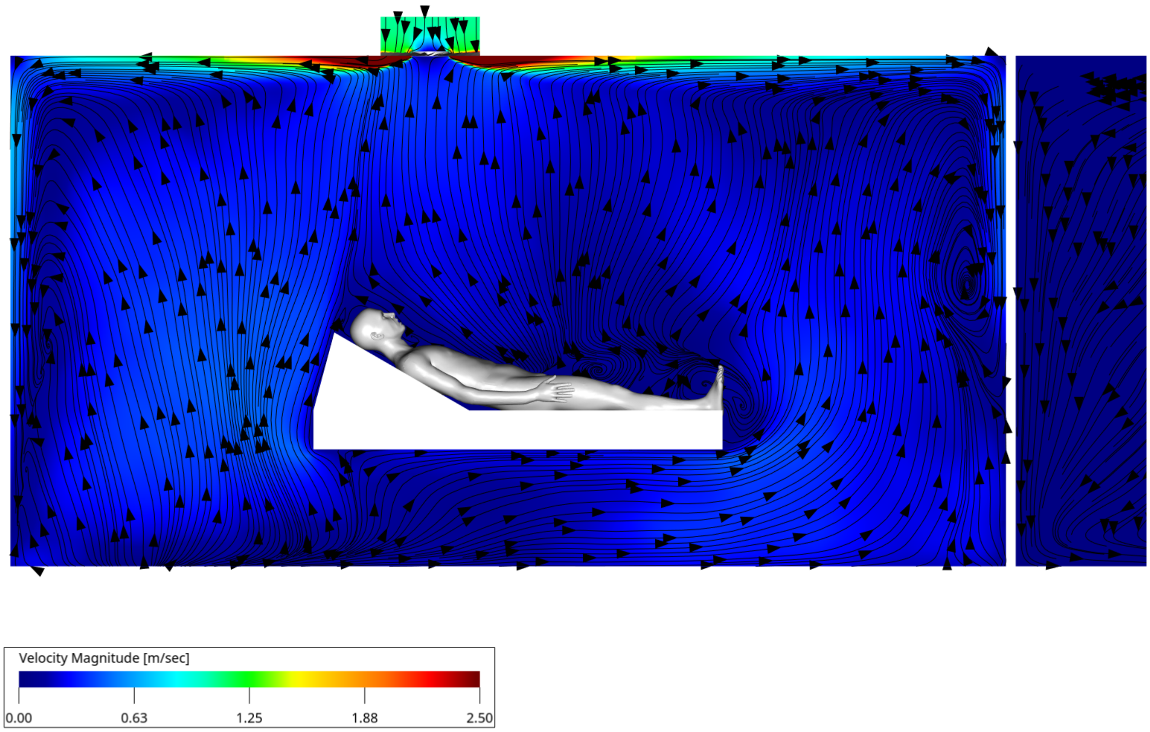


**Supplementary Fig. S8.** Simulated air velocity magnitude around the patient at Y constant position. The air flow around the patient is oriented towards the ceiling.

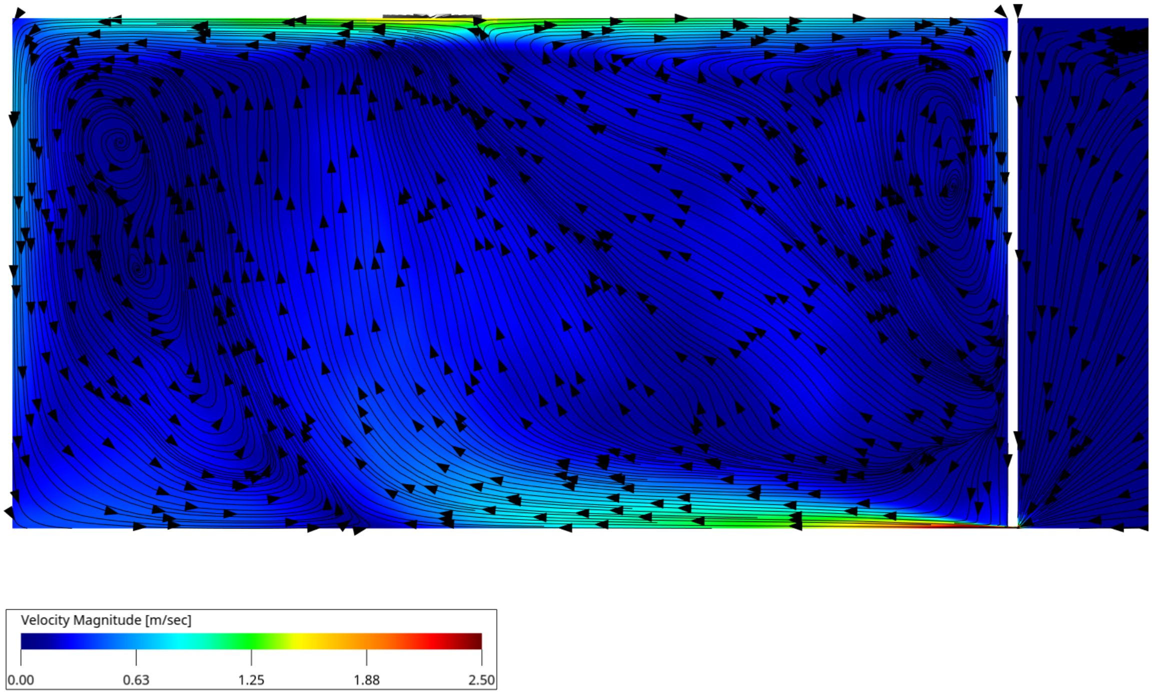


**Supplementary Fig. S9.** Simulated air velocity magnitude at the door location and Y constant position. A strong air flow comes from the gap under the door because the room is under negative pressure.

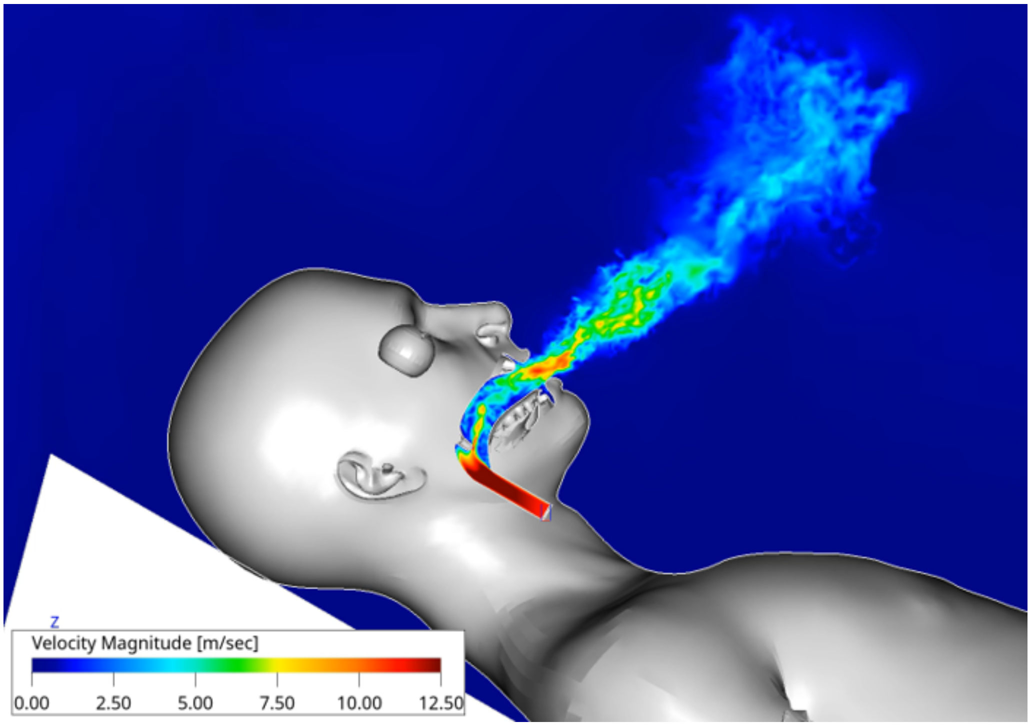


**Supplementary Fig. S10.** Instantaneous air velocity inside and outside the mouth of the virtual mannequin

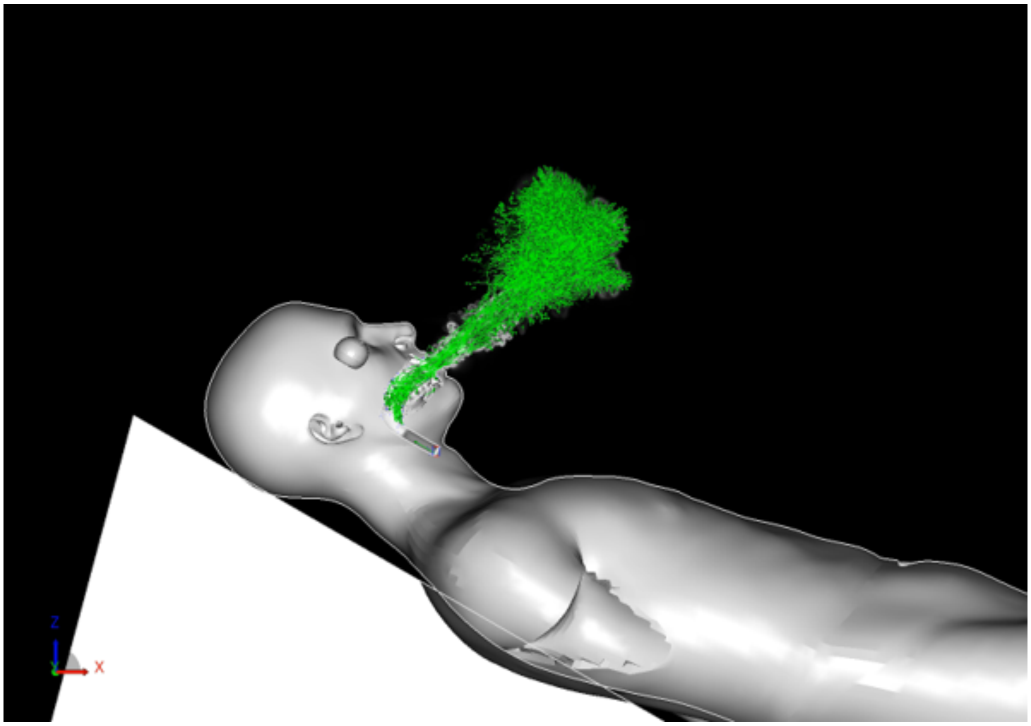


**Supplementary Fig. S11.** Particles emitted by the patient at the beginning of the simulation

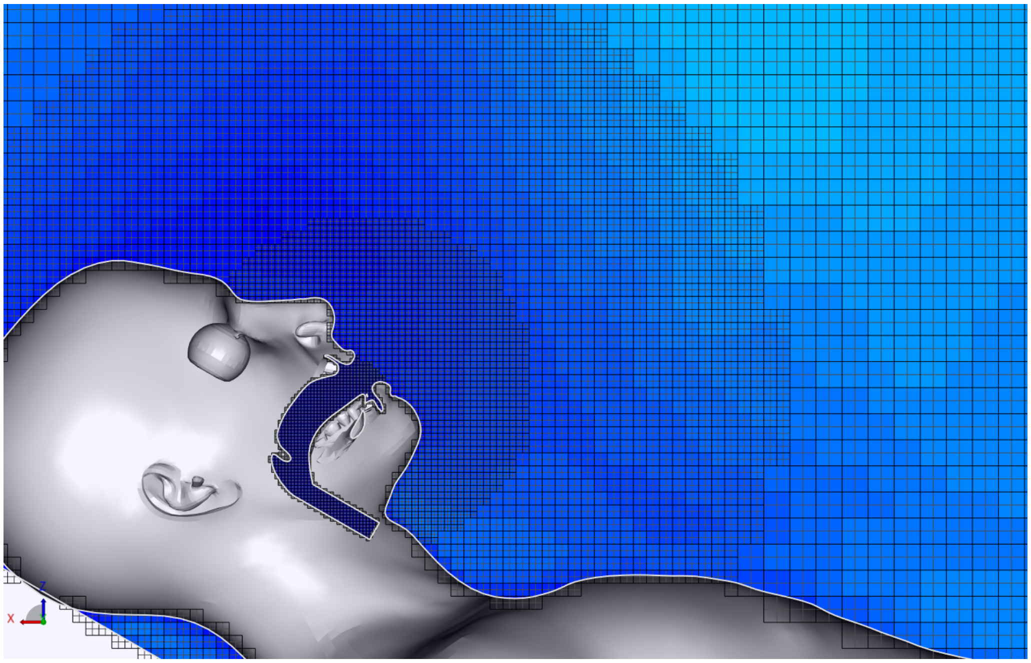

***Supplementary Fig. S12.*** *Distribution of voxels around the patient's head*

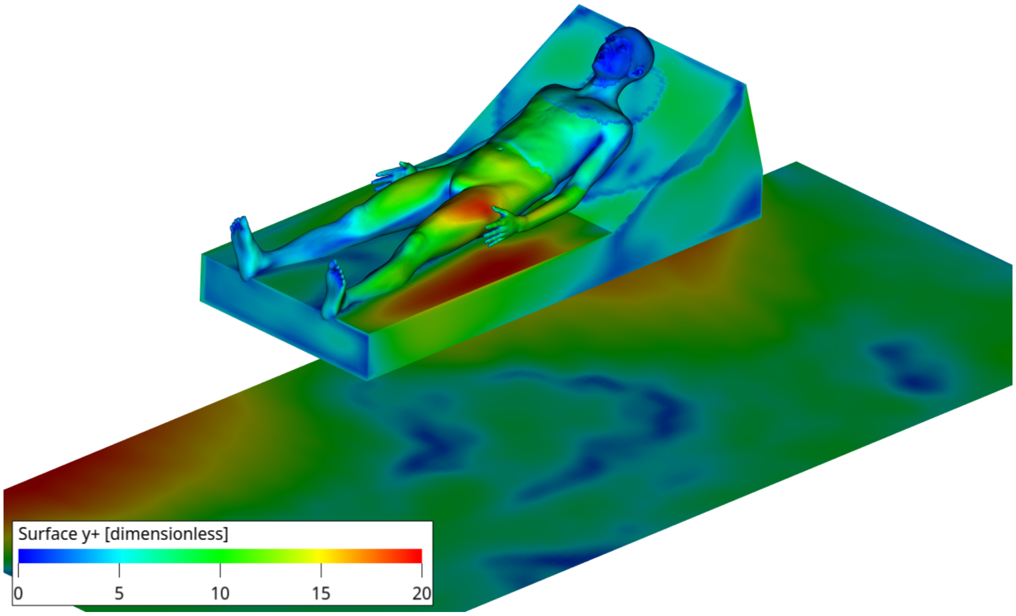


**Supplementary Fig. S13.** Y+ map around the patient

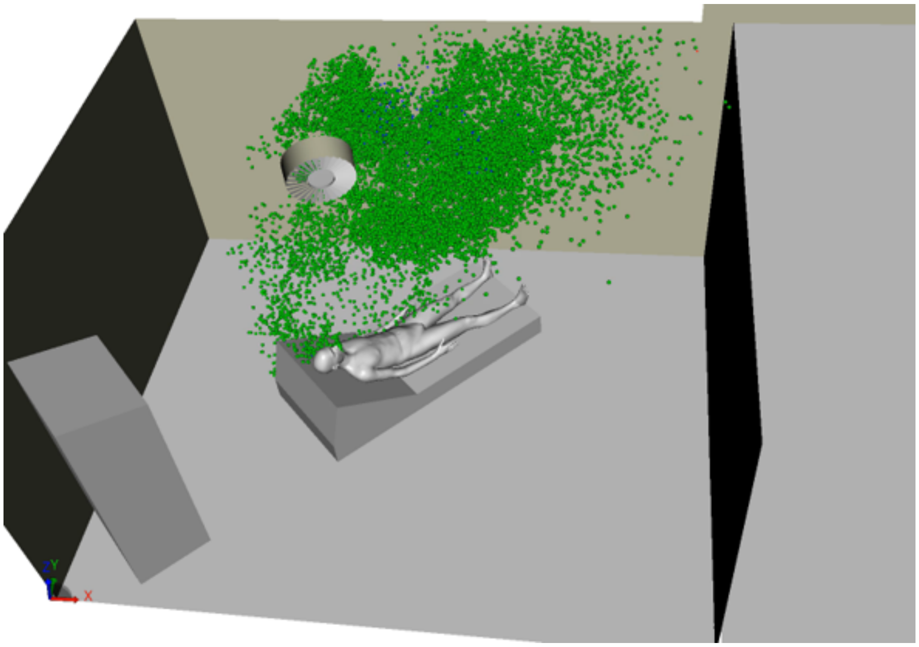


**Supplementary Fig. S14.** 3D visualization of the numerically modeled particles that are deposited at t = 45s for the first Plasmair configuration. Adding a mobile air treatment unit behind the bed helps also to reduce the amount of deposits within a room.

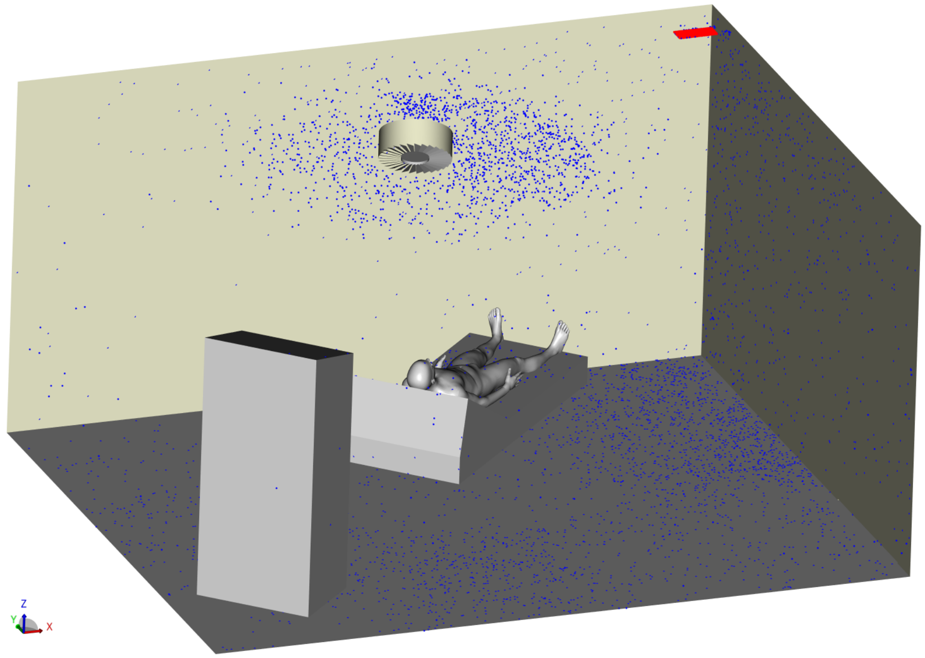

**Supplementary Fig. S15.** 3D visualization of the numerically modeled airborne particles emitted by a patient coughing in an ICU room under negative pressure for the second Plasmair configuration (t = 5s).

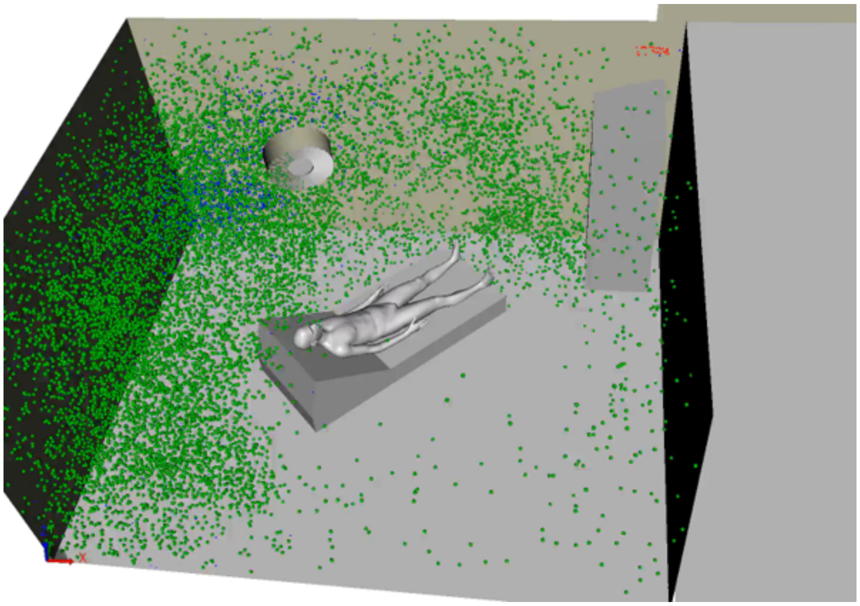

**Supplementary Fig. S16.** 3D visualization of the numerically modeled airborne particles emitted by a patient coughing in an ICU room under negative pressure for the second Plasmair configuration at t = 20s. Adding a mobile air treatment unit in front of the bed does not change anything at an early stage.

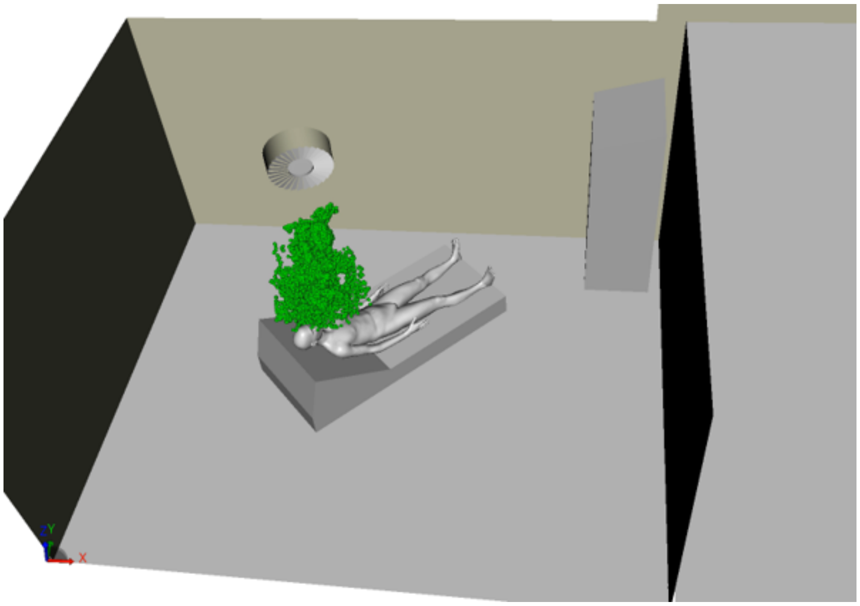

**Supplementary Fig. S17.** 3D visualization of the numerically modeled airborne particles emitted by a patient coughing in an ICU room under negative pressure for the second Plasmair configuration (t = 45s). Adding a mobile air treatment unit below the ventilation outlet increases notably the dispersion of aerosols within the room and should avoided.

**Supplementary Video S1 (separate file).** Schlieren images of a volunteer's cough with HFNC at 60L/min (View from above).

**Supplementary Video S2 (separate file).** Schlieren images of a volunteer's cough with HFNC at 60L/min (Side view).

**Supplementary Video S3 (separate file).** Schlieren images of a volunteer's cough with CPAP at 30L/min (View from above).

**SI References**


1. Gupta JK, Lin C- & Chen Q (2009) Flow dynamics and characterization of a cough. Indoor Air 19(6): 517-525.
2. Zayas G, et al (2012) Cough aerosol in healthy participants: Fundamental knowledge to optimize droplet-spread infectious respiratory disease management. BMC Pulmonary Medicine 12(1): 11.
